# A framework for integrated clinical risk assessment using population sequencing data

**DOI:** 10.1101/2021.08.12.21261563

**Authors:** James D. Fife, Tho Tran, Jackson R. Bernatchez, Keithen E. Shepard, Christopher Koch, Aniruddh P. Patel, Akl C. Fahed, Sarathbabu Krishnamurthy, Regeneron Genetics Center, DiscovEHR Collaboration, Wei Wang, Adam H. Buchanan, David J. Carey, Raghu Metpally, Amit V. Khera, Matthew Lebo, Christopher A. Cassa

## Abstract

**Importance:** Clinical risk prediction for monogenic coding variants remains challenging even in established disease genes, as variants are often so rare that epidemiological assessment is not possible. These variants are collectively common in population cohorts -- one in six individuals carries a rare variant in nine clinically actionable genes commonly used in population health screening.

**Objective:** To expand diagnostic risk assessment in genomic medicine by integrating monogenic, polygenic, and clinical risk factors, and to classify individuals who carry monogenic variants as having elevated risk or population-level risk.

**Design, Setting, and Participants:** Participants aged 40-70 years were recruited from 22 UK assessment centers from 2006 to 2010. Monogenic, polygenic, and clinical risk factors are used to generate integrated predictions of risk for carriers of rare missense variants in 200,625 individuals with exome sequencing data. Relative risks and classification thresholds are validated using 92,455 participants in the Geisinger MyCode cohort recruited from 70 US sites from 2007 onward.

**Conclusions and Relevance:** Using integrated risk predictions, we identify 18.22% of UK Biobank (UKB) participants carrying variants of uncertain significance are at elevated risk for breast cancer (BC), familial hypercholesterolemia (FH), and colorectal cancer (CRC), accounting for 2.56% of the UKB in total. These predictions are concordant with clinical outcomes: individuals classified as having high risk have substantially higher risk ratios (Risk Ratio=3.71 [3.53, 3.90] BC, RR=4.71 [4.50, 4.92] FH, RR=2.65 [2.15, 3.14] CRC, logrank p<10^-5^), findings that are validated in an independent cohort (*χ*^2^ p=9.9x10^-4^ BC, *χ*^2^ p=3.72x10^-16^ FH). Notably, we predict that 64% of UKB patients with laboratory-classified pathogenic FH variants are not at increased risk for coronary artery disease (CAD) when considering all patient and variant characteristics, and find no significant difference in CAD outcomes between these individuals and those without a monogenic disease-associated variant (logrank p=0.68). Current clinical practice guidelines discourage the disclosure of variants of uncertain significance to patients, but integrated modeling broadens this risk analysis, and identifies over 2.5-fold additional individuals who could potentially benefit from such information. This framework improves risk assessment within two similarly ascertained biobank cohorts, which may be useful in guiding preventative care and clinical management.

**Key Points:** *Question:* Can personalized risk assessments that consider monogenic, polygenic, and clinical characteristics improve diagnostic accuracy over traditional variant-level genetic assessments?

*Findings:* In established disease genes, we predict many carriers of variants of uncertain significance have significantly elevated risk. Conversely, we identify a substantial number of patients with known pathogenic coding variants who are unlikely to develop associated disorders.

*Meaning:* Many individuals would not learn about elevated risk for disease under current genetic diagnostic guidelines. Integrated risk assessments provide significant benefits over variant-only interpretation, and should be further evaluated for their potential to optimize clinical management, inform preventive care, and reduce potential harms.

## Introduction

Mapping germline variants to personalized clinical risk is a major goal in precision medicine.^1^ While diagnostic testing has advanced dramatically, clinical risk assessment remains challenging, and is generally done at the variant or gene level.^2^ To date, such genetic testing has largely been conducted in the presence of a phenotypic indication, where the prior probability of detecting a causal variant is considerable.^3^ Subsequent population screening efforts have identified substantial incomplete penetrance or reduced expressivity in these previously identified monogenic disease variants,^4–6^ and assessed how the risk attributable to these variants can be modified by clinical and polygenic risk factors.^7–9^ The UK Biobank (UKB) has generated genetic sequencing linked with clinical data, improving the ability to assess risk for individuals without clinical indication.^10^

In particular, there has been great interest in understanding the prevalence and penetrance of variation in clinically actionable disease genes.^11^ In this study, we analyze nine genes responsible for hereditary breast and ovarian cancer, Lynch syndrome, and familial hypercholesterolemia, which are designated as having sufficient evidence for population health screening by the U.S. Centers for Disease Control and Prevention^12^ and clinical actionability by the American College of Medical Genetics (ACMG).^11^ Given their established causality in clinical syndromes, many individuals have undergone diagnostic testing in these genes, revealing numerous variants meeting the classification criteria for being pathogenic or likely pathogenic (P/LP).^13^ At the population level, there is a substantial burden of such pathogenic variation: in the UKB, variants classified as P/LP within these 9 genes were identified in 0.9% of participants^14^, similar to the rate identified in prior studies.^15–17^

However, these rates do not capture the true scale of the disease burden in precision medicine, as there are likely many additional variants which confer clinical risk.^18^ There are 18-fold more rare, non-synonymous variants (population allele frequency<=0.005) observed in these genes than those already classified as P/LP, making such variants collectively common. This rate is consistent in the UKB and other large population sequencing cohorts (**Figure 1A**). Despite extensive diagnostic testing, the majority of these variants have only been observed in a few cases or controls, if at all.^3^ On the basis of their low frequencies, lack of observations in clinical cases, and/or availability of functional data, most of these variants have been classified as variants of uncertain significance (VUS). While some of these VUS may not substantially increase risk, it is unclear the extent to which moderately damaging variants contribute to clinical syndromes. A recent prospective study of cancer patients undergoing diagnostic screening identified that 47.4% carried a VUS in an established cancer gene, but only 57% of these patients met the clinical guidelines for diagnostic testing based on family history. This is suggestive that a sizable number of these variants may be incompletely penetrant if there are insufficient related individuals to meet clinical guidelines for diagnostic sequencing.^19^

**Figure 1:**
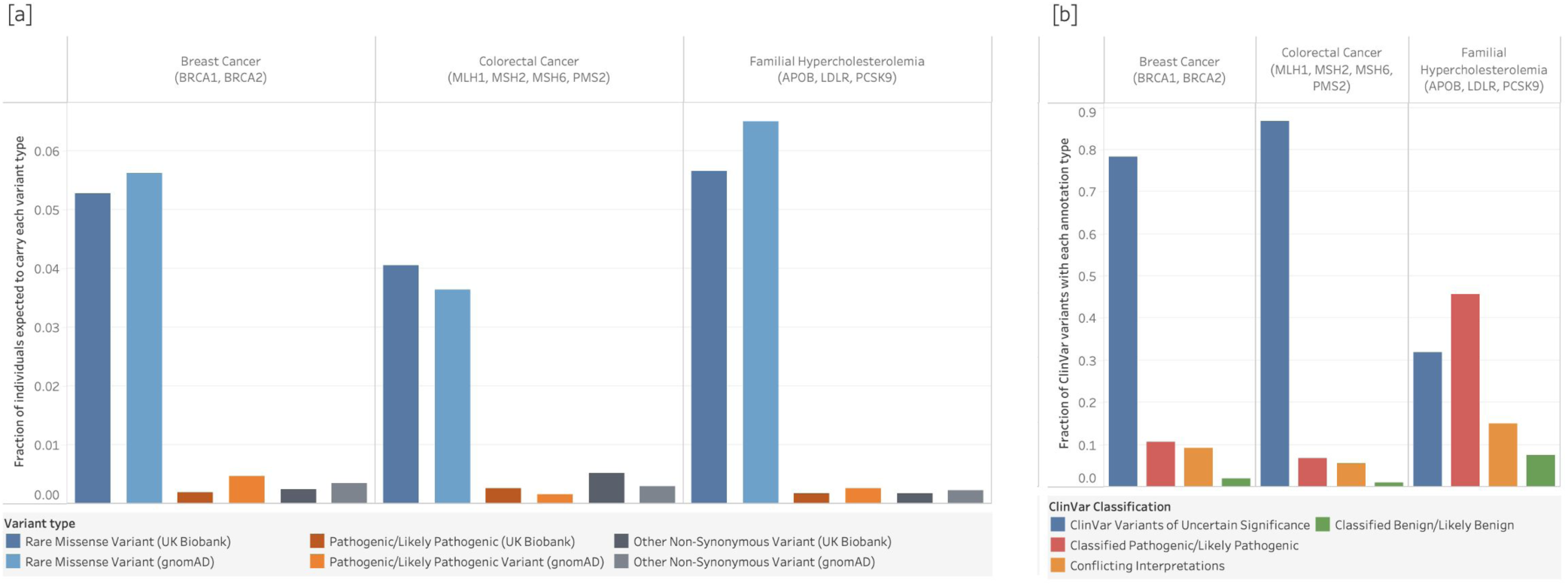
Scale of the diagnostic interpretation problem in precision medicine. **[a]** Approximately 1 in 6 individuals carries a rare, non-synonymous variant in the 9 clinically actionable genes in this study. This is unlikely to be related to dataset ascertainment as rates are similar in both the UKB (15.94%) as well as gnomAD v2.1.1 (16.64%). This is 18-fold greater than the rate of diagnostic laboratory confirmed pathogenic or likely pathogenic variants identified in the UKB dataset.^14^ Variants were restricted to non-synonymous (missense or more damaging), with allele frequency of 0.5% or lower in all population groups, and were filtered by a list of regions known to present challenges in next generation sequencing and calling (Methods). **[b]** Among variants which have been evaluated by a diagnostic laboratory and submitted to ClinVar, 68.4% are of uncertain or conflicting interpretation within the three disorders examined. These unresolved interpretations in clinically actionable genes pose challenges in clinical management and risk prediction.^2^

Among over 28,000 missense variants assessed by clinical labs in these 9 genes, 59.5% are classified as VUS and 8.9% have conflicting interpretations between P/LP and VUS, accounting for 68.4% of variants which do not have clear guidance to communicate to patients (ClinVar 6/19) (**Figure 1B**). A recent study found that fewer than 1% of these VUS were reclassified over a 3.5 year period, indicating that little headway is being made on this problem.^20^ These conflicting or uncertain assessments limit clinicians’ ability to personalize risk management recommendations and have been reported to be associated with overtreatment (*e.g.*, unnecessary surveillance or prophylactic surgery).^21^

This is exacerbated by the fact that conventional standards to assess pathogenicity are designed to provide clinical certainty at the *variant* level, without considering patient-specific predictive information that could influence disease risk, penetrance, or expressivity. This poses two challenges in the implementation of genomic medicine:

1) For most individuals who have no clinical indication for testing (say, those prospectively screened, or in biobanks), it is unlikely they will be informed about a VUS based on current practice guidelines, even if it confers high risk. Even impressively powered cohort studies focused on known disease genes cannot yet address this issue for rare VUS. A recent breast cancer analysis of over 113,000 women found that rare missense variants in *BRCA1* and *BRCA2* which have previously been classified as pathogenic confer similar risk to protein-truncating variants (odds ratios [O.R.] 16.11 and 5.68 for *BRCA1* and *BRCA2*, respectively).^22^ But in contrast, the authors found that rare missense variants (in aggregate) only confer a modest risk in *BRCA1* O.R. 1.11 (p=0.01), and no significant aggregate risk in *BRCA2* O.R. 0.98 (p=0.50).^22^ In this study, we are able to significantly discriminate risk for carriers of these rare VUS, potentially expanding the breadth of risk data that can be used to improve clinical management.

2) The current focus on variant-level interpretation leaves patient-specific predictive information (*e.g.,* sex, polygenic risk, family history) out of clinical risk assessments. Considering the magnitude with which clinical risk factors^6, 14^ and polygenic risk^9, 23^ are known to modify monogenic disease risk and contribute to penetrance of disease-causing variants, it is prudent to integrate them to distinguish between *patients* at higher and lower risk. Adjusting clinical risk assessments using these features could better inform preventive medicine and surveillance, improve genetic counseling, and even optimize therapeutic interventions.^24^

Prior studies have evaluated the relative contributions of monogenic and polygenic risk,^25^ polygenic and clinical risk factors,^26^ and clinical and monogenic risk factors.^14, 19^ Here, we develop a framework that integrates these three established classes of data to provide personalized risk assessments. Using clinical outcomes and sequencing data from the UKB, we fit models in nine disease genes that include variant-level annotations (allele frequency, computational predictive tools, protein region, and functional consequence), polygenic risk, as well as individual-level clinical risk factors (age, sex, and family history). We use these estimates to evaluate the distribution of clinical risk associated with existing diagnostic classifications (P, LP, and VUS), to classify patients who carry VUS as having higher risk or population risk, and to identify patients with known pathogenic variants who may only have population level risk. This framework provides personalized risk assessments for similarly ascertained individuals, and we make available a web application and pre-trained models to generate such predictions, as well as a software pipeline to extend this framework to cohorts beyond the UKB and our US-based biobank validation cohort.

## Results

### Creating personalized clinical risk assessments

We develop risk predictions for BC, CRC, and FH for each UKB participant who carries a rare coding variant in 9 genes related to these disorders. To do this, we fit a Cox Proportional-Hazards model^27^ separately for each gene under 10-fold cross validation to generate risk models. We then validate these models in a separately ascertained US-based biobank cohort. Prior to our evaluation, rare non-synonymous variants were classified by an ABMGG board-certified laboratory geneticist blinded to any phenotypic information according to current clinical standards for a subset of UKB participants in these genes.^2^ All risk predictions are generated for each individual blinded to this existing clinical diagnostic data. Classification guidelines, cross validation strategy, and criteria for predictions are described in the Methods. Model parameters include monogenic variant characteristics, patient-level polygenic risk, and other patient-level clinical risk factors. Model features varied in predictive power across genes (**Supplementary Table 1**), and feature selection was carried out on a per-gene basis (see Methods). Variant-level features in each model include allele frequency, measures of conservation,^28, 29^ CADD (Combined Annotation Dependent Depletion),^30^ variant functional consequence, sequence context, and protein region (see Methods).

### Evaluating the accuracy of risk predictions

We find a broad distribution of clinical risk predictions for carriers of VUS and P/LP variants, with many individuals predicted to have little or no increased risk (**Figure 2A**). Carriers of P/LP variants have significantly higher predicted risk (p<0.001 in all genes) **(Supplementary Tables 2A-B).** Additionally, we identify VUS carriers with elevated risk for each associated condition, classifying patients into higher risk (HR) and population-risk (PR) groups based on their predicted hazard. We set a classification threshold to maximize the difference in clinical outcomes among VUS carriers **(Figure 2A, Methods)**.

**Figure 2:**
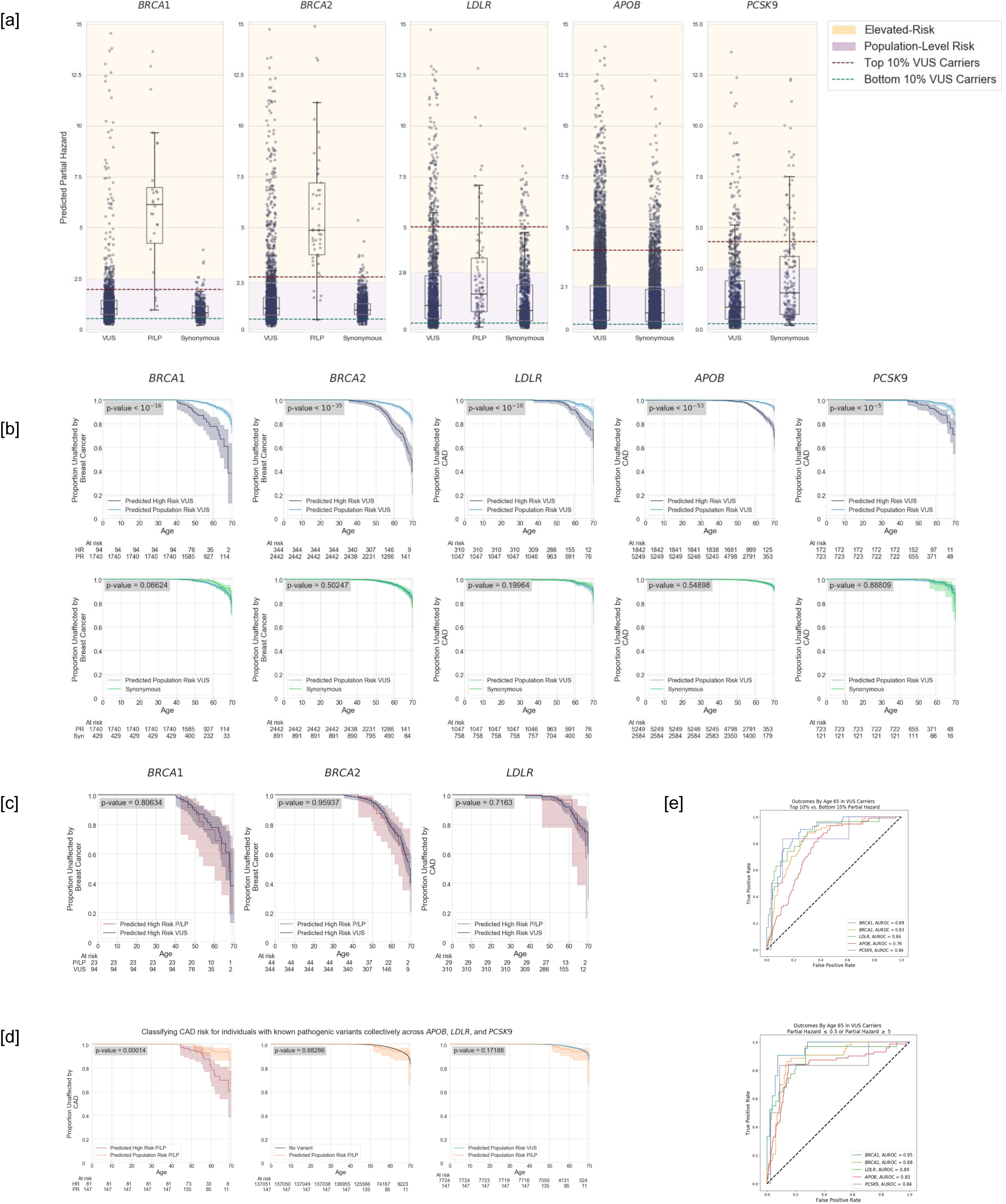
Clinical risk classifications for UK Biobank patients who carry Variants of Uncertain Significance (VUS). **[a]** Distribution of predicted risk for each gene and prior variant classification. Each point represents a hazard prediction for one patient in the UKB. High and low risk cutoffs (purple and orange shading) are determined by the maximum statistical difference in clinical outcomes (logrank p-value) among individuals above and below the threshold value. **[b]** Top row: significant differences in clinical risk are identified between patients with VUS who are predicted to have high risk versus those predicted to have population risk. Bottom row: Non-significant differences in clinical risk between VUS carriers predicted to have population risk versus carriers of synonymous variants. **[c]** We find no significant risk difference in risk between carriers of lab-classified pathogenic variants and carriers of VUS who are predicted to have high risk in genes with sufficient numbers of carriers for analysis. **[d]** Analysis of all P/LP carriers of variants in genes related to familial hypercholesterolemia (FH). We predict many individuals with lab-classified pathogenic variants to have low risk for CAD despite carrying the variant. We find significant differences in clinical risk between individuals who are predicted as higher risk (HR) vs. population risk (PR), and observe no significant difference between individuals predicted to have population risk and those who carry no variant, or VUS carriers predicted to have population risk. **[e]** Receiver operating characteristic (ROC) curves for classification of disease risk for each gene model under cross validation.

We classify 18.2% of VUS carriers as HR, accounting for 2.56% of the total population (**Supplementary Figure 3**). We find that these patients are at significantly increased lifetime risk over individuals classified as PR (logrank p<10^-5^ in each gene) (**Figure 2B**, top). These HR VUS carriers are at substantially increased clinical risk when compared to clinical outcomes of PR VUS carriers: Risk Ratio (RR) of 3.71 [3.53, 3.90] for BC (females at age 65, logrank p<10^-51^), RR of 4.71 [4.50, 4.92] (all sexes age 65, logrank p<10^-63^) for CAD, and RR of 2.65 [2.15, 3.14] (all sexes age 65, logrank p < 10^-5^) for CRC (**Supplementary Figure 2A)**. Similar risk ratios are observed between HR and PR carriers when restricting to more diagnostically challenging variants: among rare missense VUS carriers (RR=2.90 [2.60, 3.20] for BC, RR=4.52 [4.31, 4.73] for CAD, RR=2.58 [1.96, 3.21] for CRC), and when restricting to VUSs not previously reviewed by a clinical lab in ClinVar (RR=2.71 [2.26, 3.16] for BC, RR=3.75 [4.26, 3.25] for CAD, RR=3.32 [2.56, 4.07] for CRC) (**Supplementary Figure 2B-C** and **Supplementary Figure 4**).

Collectively, we classify that 81.8% of VUS carriers have PR of each associated disorder, accounting for 10.5% of the UKB population). We find that these individuals have no significant difference in risk when compared to individuals who carry synonymous variants **(Figure 2B, bottom row)**. Conversely, we find that VUS carriers predicted as HR have similar clinical risk trajectories as individuals who carry previously identified P/LP variants. In the three genes with sufficient numbers of individuals with pathogenic variants, P/LP carriers predicted as HR do not have significantly different clinical outcomes from VUS carriers predicted as HR (**Figure 2C**).

Consistent with diagnostic classifications, we find only a small proportion of women with known pathogenic BC variants who are classified as having PR (*N=*5 *BRCA1* (18%) and N=5 *BRCA2* (10%)). However, among the 231 individuals with known pathogenic variants in *APOB*, *LDLR*, and *PCSK9,* we find a substantial number of individuals with conflicting integrated risk classifications compared with variant-only diagnostic classifications. For 84 individuals predicted as HR, we identify a significantly increased risk of CAD than in the P/LP carriers classified as PR (RR=3.70 [2.92, 4.48], logrank p=1.4x10^-4^). **Notably, 64% of individuals who carry known pathogenic FH variants are not predicted to have high risk of CAD**. We find no significant difference in CAD risk between these 147 individuals (N=147/231) compared with individuals with no variant in these genes (logrank p=0.683) or when compared to PR VUS carriers (logrank p=0.172) (**Figure 2D**). In CRC, among the 45 carriers of P/LP variants, we predict 31 will have PR (59.2%) (logrank p=0.008). We also find high measures of classification accuracy when predicting clinical outcomes of individuals with very high or low risk (*AUROC* ∈ [0.76, 0.95]) (Figure 2E).

We provide a web application that generates personalized estimates of lifetime and age-specific risk for variant carriers in these nine genes (https://clintegrate.herokuapp.com/), as well as a downloadable python package to generate estimates of clinical risk (https://github.com/j-fife/clintegrate/).

### Validation of risk classification in a second, independent biobank

We validate our risk predictions in an independent set of 92,455 individuals from the MyCode biobank in the US-based Geisinger Health System (Methods). Using the predictive models generated for each gene from the UKB dataset, we estimate the clinical risk for each patient carrying a non-synonymous coding variant in the three FH and two BC genes previously evaluated. We find statistically significant differences in predicted risk among individuals developing or not developing BC or CAD by age 65 (p ≤ 0.011) (**Figure 3A**). Patients who are predicted as HR are also significantly enriched in early onset phenotypes (*χ*^2^ p=9.9x10^-4^ for BC, *χ*^2^ p=3.72x10^-16^ for CAD) with large differences in risk among carriers predicted as HR versus PR (RR=3.92 [3.18, 4.66] for BC, RR=2.39 [2.18, 2.6] for CAD) (**Figure 3B**). We also find similar classification accuracy for predicting clinical outcomes (*AUC* ∈ [0.68, 0.92]) (**Figure 3C**). Though there are limited numbers of novel variants in the MyCode cohort which are not already observed in the UKB, we identify significant differences when restricting to novel variant carriers in many genes (**Supplementary Table 5**).

**Figure 3:**
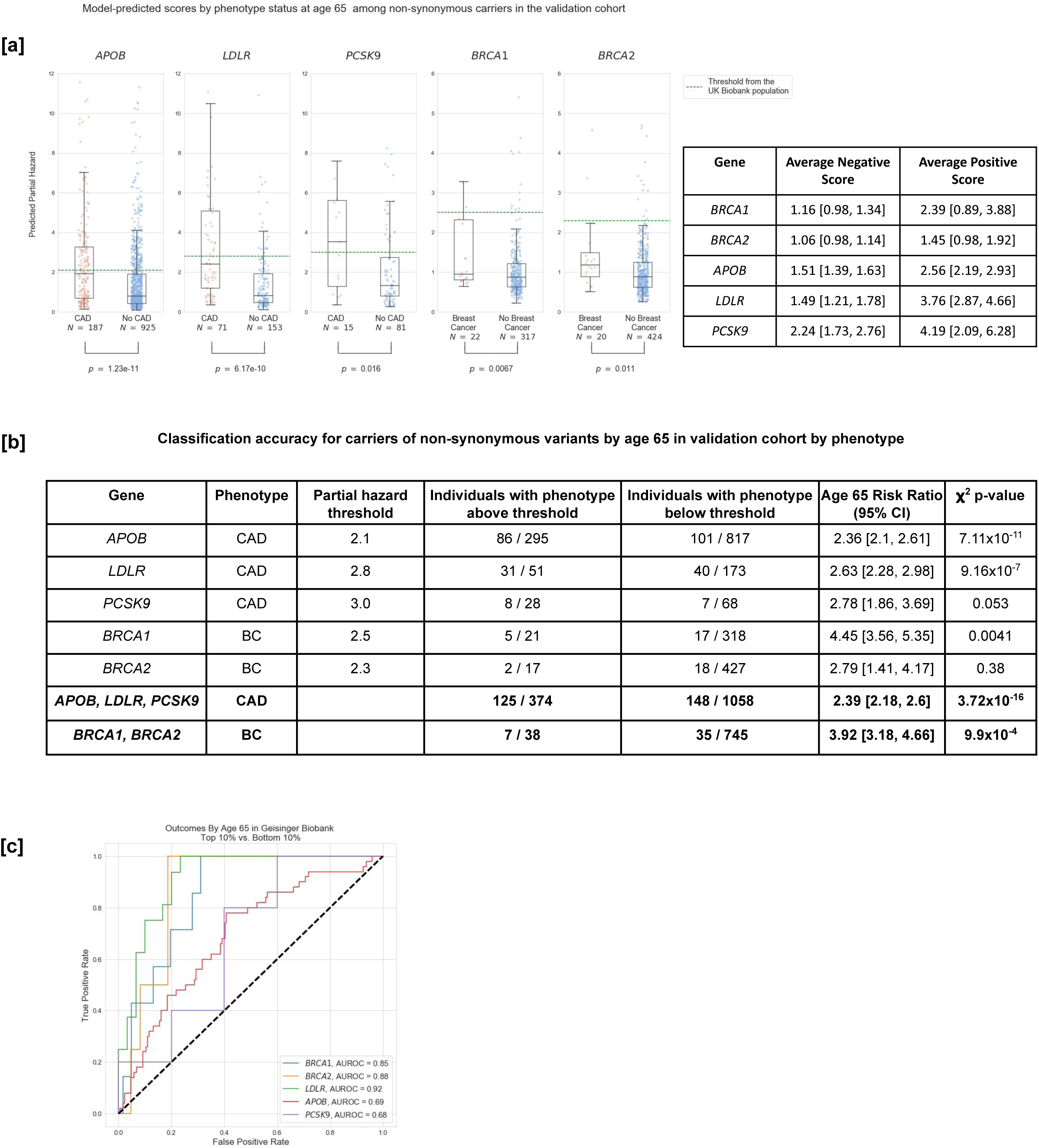
Validation of predicted risk in a second biobank (Geisinger MyCode cohort, N=92,455 individuals). **[a]** Gene-based predictive models were fit using UKB participant data, and used to make estimates of clinical risk for individuals in the validation cohort. We find that individuals with early-onset coronary artery disease (CAD) or breast cancer (BC, females before age 65) have significantly higher predicted hazards than those unaffected by age 65 (Mann-Whitney U). **[b]** We find significant enrichment of early-onset CAD or BC cases in the validation cohort as determined by *χ*^2^ p-value, among individuals predicted to have high risk by the UKB model by gene, and by phenotype. **[c]** Receiver operating characteristic (ROC) curves for disease classification in the validation cohort using scores generated by UKB gene risk models.

### Defining protein regions that differentiate clinical risk

Variants that are within protein domains^31, 32^ and in proximity in the folded protein^33^ to pathogenic variants are expected to be under increased negative selection. Accordingly, proximity to such variation may also confer increased clinical risk,^34^ and is considered moderate evidence in support of pathogenicity in clinical assessment.^2^ Conversely, there may also be “cold spots,” where most variation is expected to be neutral^35^ or other regions where mutations may confer protective effects.^36^ Given the broad spectrum of potential impacts of missense variants and the utility of proximal variation, we sought to define regional risk to quantitatively discriminate between regions with missense variants that confer higher or lower risk. Our risk predictor uses a novel approach to model missense variant risk by partitioning each protein into regions.

To define regions of differential clinical risk, we make use of the abundant clinical diagnostic data in these genes from ClinVar, curated independently from the UKB. We filter to known pathogenic single nucleotide variants which would be expected to impact a specific region rather than have a downstream effect (*e.g.,* non-terminating, non-splice, non-frameshift variants, see Methods). We use Jenks natural breaks one-dimensional optimization^37^ to partition the protein into regions with differential rates of pathogenic reports, with specific nucleotide breakpoints. The number of regions is pre-defined per 1kb coding sequence length (*e.g 13* regions for *APOB*: 13,689bp, 5 regions for *BRCA1*: 5,589bp) with a minimum of 5 regions (**Figure 4A**). Regions without adequate sequence coverage are treated as missing.

**Figure 4:**
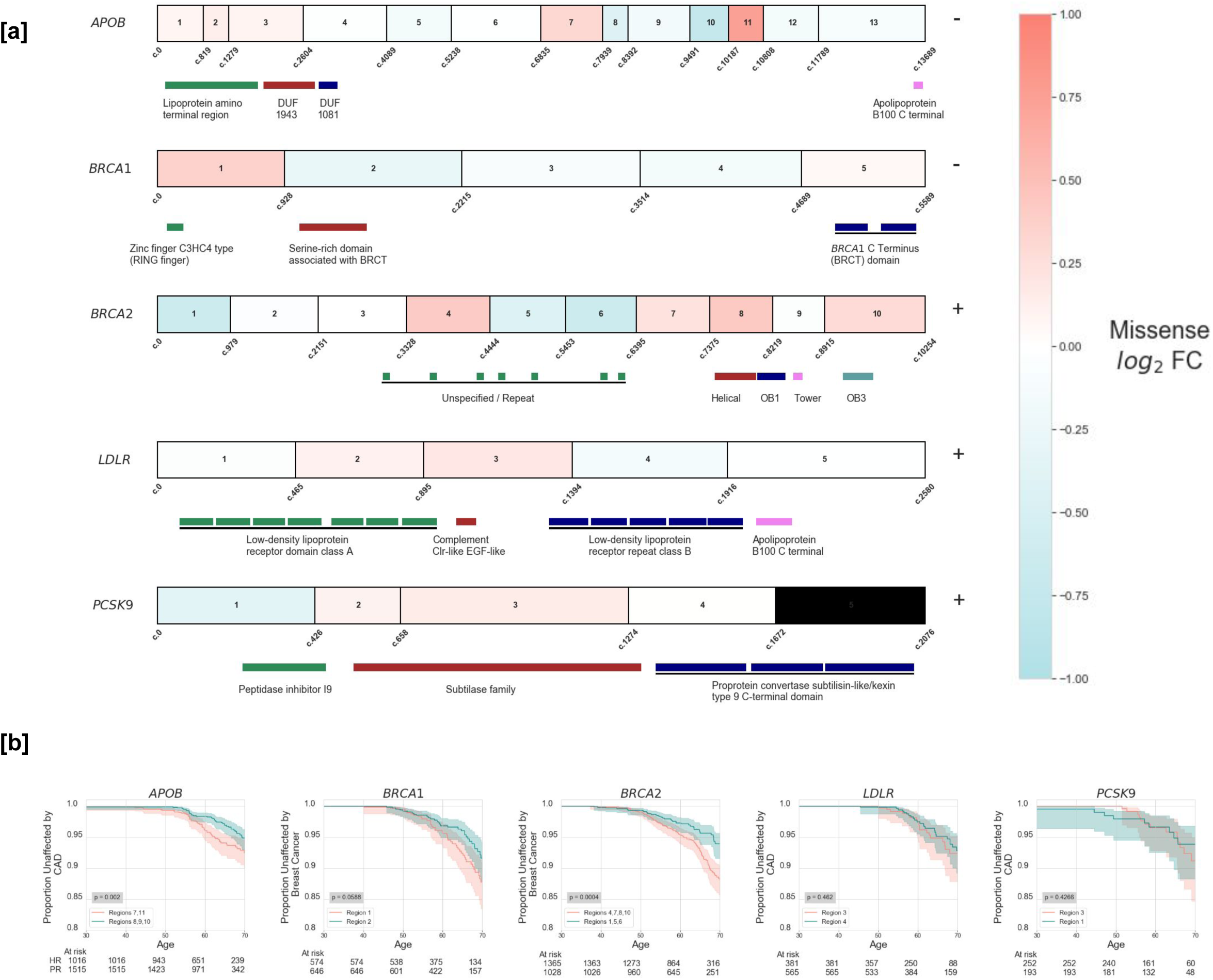
Defining regions within each protein with differing clinical risk. We identify regions in each gene that are associated with higher and lower risk for breast cancer (BC) and coronary artery disease (CAD) using previously identified pathogenic variants (ClinVar) without the use of UKB patient data. **[a]** Using previously identified pathogenic variation from ClinVar, we use Jenks natural breaks optimization to partition each protein into regions (Methods). Hazard ratios are calculated for missense variants in each region, and the log_2_ fold change (FC) from the overall gene missense hazard ratio is listed by region. *PCSK9* has one section with missing sequencing data (black). **[b]** Kaplan-Meier estimates (95% confidence intervals) for individuals with missense variants in regions defined as higher risk (red) or lower risk (blue) (logrank p-values). In genes where no variant met either the high or low-risk classification, the region with the highest or lowest risk was used in the comparison.

We establish a baseline hazard for all carriers of missense variants, regardless of coding position. The risk in each region is then compared to the baseline risk by log_2_ fold change (L2FC) (**Figure 4A**). We consider segments with L2FC over 1.1 to be higher-risk regions (HRR) and regions with L2FC under 0.9 to be lower-risk regions (LRR). We find at least one region of elevated or decreased risk in all five genes analyzed (**Supplementary Figure 5**). Further, we observe significant differences in clinical outcomes when comparing individuals with missense variants in HRRs versus LRRs (**Figure 4B**). Inclusion of regional data improves model concordance and log-likelihood in all genes analyzed (**Supplementary Table 3**).

In *BRCA2,* individuals with missense variants in HRRs develop breast cancer at a higher rate than those with missense variants in LRRs (RR=1.68 [1.28, 2.07], logrank p=4.0x10^-4^). These regions may be particularly informative in genes where missense variants have different functional effects (*e.g.,* gain vs loss-of-function). In *APOB*, we recover known pathogenic variants in exon 26 which increase risk (Region 11) and regions where LoF variants would be expected to lower risk (Regions 8-10) (RR=1.97 [1.53, 2.41], logrank p=0.0020, all HRRs vs LRRs).^38^ We are also able to differentiate risk even in a gene where most variation lowers risk — we find four additional regions which confer higher missense risk than average in *APOB,* improving discrimination of missense risk (Regions 1-3 and 7) (Age 60 RR = 1.36 [1.14, 1.58] logrank p=0.044). This *APOB* finding is replicated in the Geisinger MyCode validation cohort when comparing missense carriers in these regions to those without coding variants (Age 60 RR = 1.28 [1.01, 1.56] *χ*^2^ p=0.044).

### Comparison to known protein domains and functional assays

Consistent with prior work, we find that our regions of increased risk are broader than protein domains.^39^ These regions provide risk estimates for larger portions of each gene (**Supplementary Figure 6A**), and consequently for substantially more variant carriers (**Supplementary Table 4**). One example of clinical utility is in *BRCA2* carriers: consistent with a prior breast cancer cohort study,^22^ we find no significant difference in clinical risk between carriers of rare missense VUS within versus outside of known protein domain boundaries (Age 65 RR=1.22 [0.86, 1.57]) (**Supplementary Figure 6B**). However, we find that the risk ratio conferred by our HRRs vs LRRs is stronger and more significant (RR=1.68 [1.28, 2.07], logrank p=0.0040). Additionally, this approach has the benefit of providing risk estimates for all variant carriers, where protein domains may only provide predictions for small numbers of variant carriers (e.g. 12 *BRCA1* RING domain carriers, with 2 developing breast cancer by age 65) (**Supplementary Figure 6E**).

Next, we compare integrated predictions to high-throughput *in vitro* assays which measure variant functional effects within *BRCA1* and *BRCA2*, which are correlated with clinical diagnostic assessments.^40, 41^ However, in individuals who carry a variant with a functional assay score, we find no significant difference in functional scores between women who do or do not develop breast cancer by age 65 in either gene (one-sided KS p=0.71 for *BRCA1*, p=0.23 for *BRCA2*). Further, we find that these scores do not outperform the predictions we assign to the same carriers (**Supplementary Figure 6F**).

## Discussion

While there are many genes with established disease associations, it is still challenging to assess the clinical risk for asymptomatic, prospectively screened patients who carry variants in them.^42^ Patients without an indication for testing have substantially different risk profiles which cannot be accurately modeled by previously described pathogenic clinical variation. We have developed a framework to provide integrated risk estimates for patients who carry germline variants. We use population sequencing data to fit models which include variant-level predictive features of monogenic variants, as well as polygenic risk and other clinical risk factors. These predictions are highly concordant with clinical outcomes in both the UKB (under cross-validation) as well as in a large independent validation cohort from Geisinger Health System.

These findings suggest that restricting to variant-level data in clinical risk assessments may be limiting the full potential of genomic medicine. Based on the inherent complexities in variant interpretation and the potential for harm from inaccurate interpretations, the ACMG and the Association for Molecular Pathology have provided guidelines which focus on classification at the variant level.^2^ These and other guidelines related to interpretation^43^ and reporting of secondary findings^44^ discourage the disclosure of VUS in asymptomatic individuals on the basis of their limited evidence, placing the disclosure emphasis on previously identified pathogenic variants in prospectively screened patients.

Here, we are able to identify individuals who carry previously unclassified variants who have significantly increased clinical risk, which could be used to improve clinical management. For example, there are 11 UKB patients who carry *LDLR* p.Arg78His, a variant with conflicting diagnostic interpretations in ClinVar (reported as benign by 1 lab, VUS by 2, and LP by 1) who have either reached 65 or otherwise developed CAD. Collectively, these participants have only a modestly elevated LDL-C (avg. 154.6). Four of these participants developed CAD even though these patients have normal LDL-C levels (avg. 146.3, 56th percentile). However, they have additional risk factors - 3 are male, 3 have a family history of CAD, and the four have a high PRS (avg. 1.36, 90th percentile) - compared to the remaining 7 carriers who have not developed CAD - only 1 is male, only 1 has a family history of CAD, and they collectively have a PRS in the 62nd percentile.

Conversely, we also find individuals with known pathogenic variants who do not have elevated risk of developing a disorder. Among patients who carry *LDLR* p.Asp472Tyr, a variant described as P/LP by 8 diagnostic labs in ClinVar and in the hydrophobic core of the protein, there are 4 carriers in the UKB who have reached age 65 with high LDL-C values (avg. 209.4, 96th percentile); however none develop CAD by age 65. This is likely on the basis of their other personal risk factors, as the group includes 4 women, none of whom have a family history of CAD, and do not have an elevated PRS (avg. 0.26, 53rd percentile of scores), The variant is also in the region of *LDLR* which we estimate to have the lowest risk (Region 4). On the basis of these factors, these individuals are predicted to have population-level risk (avg. 0.52, 30th percentile).

When patients and clinicians have incomplete or incorrect information, it can result in serious clinical and/or sociological consequences, either through unnecessary surveillance or irreversible prophylactic surgery.^45, 46^ In one retrospective study of *BRCA1* or *BRCA2* VUS carriers, 39% of individuals without cancer opted to receive a bilateral mastectomy, and among all patients with and without cancer, and 21 of 97 or 22% of patients had a VUS reclassified, with 95% downgraded to benign.^47^

Our findings suggest that integrated risk predictions are sufficiently informative to warrant prospective evaluation in a clinically realistic setting. It is vital to measure the risks and benefits of increased surveillance, or changes to clinical management compared with existing standard of care. The genes in this study are increasingly being sequenced in biobanks and reviewed in patients to identify secondary findings, so there is a natural population of P/LP carriers who might benefit from additional personalized context of reduced risk, or VUS carriers who might benefit from a broader consideration of their risk factors.

By building each model by gene, we fit the unique regional context and risk profile for variants in each protein, and recapitulate known protein regions harboring increased risk. For example, in *BRCA1* while missense VUS collectively confer little to no increased clinical risk, the regions we specify as higher risk contain the RING and BRCT domains which contribute to ubiquitin ligase activity, cell cycle control, where variation is known to increase cancer predisposition.^48, 49^ These protein regions are part of the hydrophobic core of *BRCA1*, while the remaining regions have few predicted structural features.^50^ As biobank sequencing cohorts expand, we expect our resolution to improve for regional estimates, and to have sufficient statistical power to model interactions between monogenic and polygenic risk.

There are several limitations to this study. While we provide direct estimates of clinical risk to enable rapid translation into clinical care, these estimates are developed using patients who are ascertained from biobanks and specific population groups. These risk estimates may not be applicable for patients who are ascertained differently or from different population groups. Importantly, we note that previous studies of polygenic risk have found limitations to the applicability of polygenic risk estimates in non-European populations,^51, 52^ indicating the urgent need to expand study cohorts and to develop population specific scores and improved trans-ethnic risk prediction methods as has recently been done with lipid levels.^53^ Clinical translation of these results will rely on broader validation and evaluation of these methods in additional population groups. Additionally, the biobanks in this study may have biases due to ascertainment or censoring.

Developing these estimates in broader numbers of genes and disorders will also require careful evaluation of monogenic, polygenic, and clinical risk factors. Future work should integrate additional established behavioral and clinical risk factors for each syndrome. Larger panels of genes will require sufficient prevalence of each disorder attributable to each gene, validated polygenic risk scores, and other individual-level predictive features. As population sequencing efforts expand, it is critical to further improve and personalize predictions of clinical risk. This is particularly important in asymptomatic individuals with no clinical risk factors or family history, as many variants in known disease genes may not confer risk or be completely penetrant. Biobanks and population cohorts are now sufficiently powered to advance genomic medicine through the development of standards that optimize surveillance, counseling, and therapeutic interventions.

## Methods

### Study design, setting and participants

The UKB is a prospective cohort of over 500,000 individuals recruited between 2006 and 2010 of ages 40-69 years.^10^ 200,625 participants with exome sequencing data were included in this analysis. Analysis of the UKB data was approved by the Mass General Brigham Institutional Review Board (Protocol 2013P001840). Work was performed under UKB application #7089. Family history of disease in parents and siblings was reported by participants using a structured assessment tool at the time of enrollment. Prediction thresholds and relative risks are validated using 92,455 individuals from the Geisinger MyCode cohort who enrolled after 2007 at over 70 recruitment sites in central and northeastern Pennsylvania^54^. From participant survey data, we identified individuals with a family history of three common, related phenotypes (reported as heart disease for FH genes, breast cancer for HBOC genes, and bowel cancer for Lynch syndrome genes). Adjustments were not made for family size. LDL cholesterol concentrations were assessed at the time of enrollment as part of the study protocol. To estimate untreated values for LDL cholesterol, measured values for participants reporting use of lipid-lowering medications were adjusted in accordance with average impact on LDL cholesterol levels, as performed previously.^23^ Polygenic risk was calculated for each patient using previously validated scores for each of the three disorders,^55–57^ using ancestry-corrected reference distributions for each score.^58^

### Exome sequencing and variant annotation

Exome sequencing was performed for UKB participants as previously described.^10^ Variant allele frequencies were estimated from the Genome Aggregation Database (gnomAD v2.1 exomes N=125,748). Variants were included with population maximum allele frequencies of <=0.001 (N=9,042) (Ensembl gnomAD plugin)^59^ or if not present in gnomAD (N=6,715). The canonical functional consequence of each variant was calculated using Variant Effect Predictor (v99).^60^ Additionally, GERP, PhyloP, population allele frequencies, and CADD scores (Ensembl REST API v15.0)^59^ were included for each variant where available, as well as variant CpG and transition/transversion status.^61^ From VEP, “CADD_raw”, “GERP++_rs”, and “phyloP100way_vertebrate” scores are listed as CADD, GERP, and phyloP in the manuscript. CpG context was obtained from PolyPhen-2 v2.2.3.^62^ Variants which are non-PASS quality in gnomAD were excluded, as well as any variants in low complexity regions, segmental duplications, or other regions known to be challenging for next generation sequencing alignment or calling.^63^ Variant functional consequences are calculated for the canonical transcript, and include synonymous, missense, or predicted LOF variants. The predicted LOF category includes frameshift, stop gained, start gained, start lost, and canonical splice-site variants. Non-coding variants outside of essential splice sites were not considered in the analysis.

### Clinical diagnostic assessment of variants

A board-certified laboratory geneticist (blinded to any patient phenotypic information) classified the pathogenicity of observed variants in the study genes according to current clinical standards. The nine study genes included three related to familial hypercholesterolemia (*APOB*, *LDLR*, and *PCSK9*), two related to breast cancer (*BRCA1* and *BRCA2*), and four related to Lynch syndrome (*MLH1, MSH2, MSH6,* and *PMS2*). Clinical variant assessments were made for all rare, non-synonymous variants (gnomAD population maximum allele frequency <=0.001), following the ACMG/AMP guidelines for variant assessment and best practices.^2^ All variants meeting these criteria which had not been previously classified as P/LP/LB/B were classified as VUS for this analysis. For P/LP variants we use annotations from the Mass General Brigham Lab for Molecular Medicine.

Individuals with multiple variants in the same gene with different annotations of P/LP/VUS were always included in the most severe category in descending order: P, LP, and VUS. Synonymous variants were grouped separately, and not labeled VUS.

### Clinical endpoints

The primary clinical endpoints were specific to each condition -- coronary artery disease (CAD) for familial hypercholesterolemia, breast cancer (BC) for HBOC syndrome, and colorectal cancer for Lynch syndrome. Case definitions for CAD, BC, and colorectal cancer were defined in the UKB using a combination of self-reported data confirmed by trained healthcare professionals, hospitalization records, and national procedural, cancer, and death registries, previously described at the disorder level.^14^ Estimated untreated levels obtained using adjustments for lipid-lowering therapies were used in analyses, as described previously.^23^

### Predictive models and feature selection for each gene

The age-dependent hazards for each disease were generated by comparing individuals affected by each condition to those unaffected, using a Cox proportional-hazard model for each gene and condition. Each model includes patient genome-wide polygenic risk for each associated disorder, previously shown to have utility in risk stratification^55^ and in assessing the variable penetrance or expressivity of monogenic variants.^64^ Though non-additive interactions between PRS and monogenic variants have been previously identified in these phenotypes in cohort studies,^25, 65^ a previous analysis in the UKB did not find significant interactions in *BRCA1* and *BRCA2*, on the basis of limited statistical power to detect it.^9^ Accordingly, these terms were not included in the model, but could be added to gene-based models fit on sufficiently powered cohorts. Clinical risk factors included age, sex, and family history derived from participant survey data, previously generated and validated in the UKB.^14^ Covariates include genetic principal components 1-4, CADD, gnomAD population maximum allele frequency, GERP, phyloP, transition/transversion, CpG/non-CpG context, variant functional consequence (synonymous, missense, predicted LOF), sex, family history, and polygenic risk score, with the predictive model standardized to the average of each of the covariates across all individuals as the baseline prediction. Statistical analyses were performed using the python lifelines survival analysis package (v0.23.9).^66^ Hazard predictions for individuals are generated using a 10-fold cross-validation process, where predictions are made for each 10% by training a separate Cox model using the remaining 90% of the participants.

A predictive model was developed for each gene separately. Features were included if univariate regression was statistically significant (p < 0.05) as a predictor for each phenotype, or if inclusion of the feature resulted in an improvement in model concordance of at least 0.01 when added to a model with all univariate significant features. Additionally, categorical variables related to functional consequence were always included (synonymous, missense, predicted LoF).

For individuals with more than one variant in a gene, the variant with the most severe functional annotation is analyzed. If this led to a patient carrying two variants of the same functional category, the variant labeled P/LP through LMM annotation was prioritized when applicable. If a patient carried multiple variants with the same functional consequence and neither had P/LP classification, the variant with the highest CADD score is included for analysis. If variants have the same CADD score, the variant with the lowest allele frequency is selected. Patients are included in each model regardless of whether they carry a variant in each gene to establish a baseline for incidence excluding variant-level factors. Individuals without variants in each gene have all variant-level features either assigned to zero or to the least severe value for each feature.

For each gene’s predictive model we removed all individuals with coding variants in other genes related to the same condition. For the *BRCA1* and *BRCA2* models examining risk for breast cancer, males were excluded from all analyses. Individuals with missing information regarding age, age of onset, or presence of the condition were not included in any analyses.

### VUS and P/LP comparisons by gene

In selecting genes to compare across annotation classes, we restrict to genes with more than 15 cases of the disorder in individuals with variants in the gene, and more than 5 unique variants designated as P/LP observed in the patient set after patient and variant filtering described previously. In *APOB* only one variant, and in *PCSK9* only three variants met our criteria. These genes were not compared in certain parts of Figure 4, but their comparisons can be found in Supplementary Figure 7. In all survival analyses related to outcomes across classes (Figure 2), we excluded individuals over age 70, and excluded individuals with coding variants in other genes related to the same disorder. In Figure 2D we use a cutoff of 3.0 predicted partial hazard when designating population-risk VUS carriers. This was due to the abundance of carriers in *APOB* when compared to *LDLR and PCSK9. APOB* has a much lower threshold, skewing the overall risk-makeup of the VUS group.

### Filtering variants used in regional boundary generation

To generate protein boundaries for differential risk stratification, we partitioned each gene restricted to pathogenic variants in ClinVar. Single nucleotide missense variants were included for analysis from ClinVar (June, 2019) which were non-canonical-splice affecting and non-terminating. Only pathogenic and likely pathogenic variants were included which had no conflicting interpretations. Reports of pathogenicity were all treated as a positional input (*e.g.,* a variant with 3 reports of pathogenicity were included as three values for the same position).

Regions were defined based on the result of Jenks natural breaks optimization for varying numbers of sections. We set the number of regions based on the length of the coding sequence as the floor of one per thousand base pairs (*e.g.,* 13 regions for *APOB*: 13,689bp, 5 regions for *BRCA1*: 5,589bp). For genes with fewer than 5,000bp we set a minimum number of 5 regions. In *PCSK9* a portion of the coding sequence was not covered in the exome sequencing data. We treat this protein region as missing, and separate the remaining 1,672bp into four regions using the Jenks optimization method (black section, **Figure 2A**). Four of the nine genes (*MLH1*, *MSH2*, *MSH6*, *PMS2*) did not have sufficient individuals with colorectal cancer and missense variants in the UKB to reliably determine differences in risk by region.

For comparisons of regions with increased risk versus those with decreased risk, we first compared the highest risk region to the lowest risk region and measured whether there was a significant difference in clinical risk. If not, the next highest- or lowest-risk regions were added to the high- or low- risk regions to optimize logrank p-value for separating clinical risk, or until all regions were exhausted. For all genes evaluated, the optimal logrank p-value is reported.

### Statistical Tests

We use the logrank test when determining differences in clinical risk for each disorder among different groups in the UKB due to the availability of age of onset in addition to participants’ current age. When measuring differences in clinical risk in the validation cohort, the chi-squared test at a specific age is used, as age of onset was unavailable. While risk plots include participants through age 70, data for these statistical comparisons was not age restricted. All risk ratios (RR) are taken at age 65 unless otherwise specified. All confidence intervals reported are 95%. Logrank p-values are generated using the python package lifelines v0.23.9.^66^ Chi square and Mann-Whitney U tests were performed using scipy v1.4.1.^67^ Reported Mann-Whitney U p-values are two-sided. Additional data processing, computation, and statistical tests were performed using NumPy v1.17.3^68^ and pandas v1.1.5.^69^ Figures were made using Matplotlib v3.1.1^70^ and seaborn v0.9.0.^71^ Box plots show the median, and quartile ranges as default parameters to seaborn, data determined by seaborn’s method as outliers are not shown on boxplots but are shown in the underlying plots of points.

### Criteria For Geisinger Biobank Variant Filtering and Individual-level features

Similar to filtering on UKB participants, when evaluating a gene’s predictive model we restrict it to individuals without non-synonymous coding variants in the other genes related to the condition. For individuals with multiple non-synonymous coding variants in the same gene, values for the variant included in the model are selected in the same fashion as selecting variants among UKB participants. As expected based on clinical ascertainment, we found differences in disorder incidence between Geisinger Biobank and UKB, and no adjustment for incidence was done when making predictions on Geisinger Biobank participants (**Supplementary Table 6**).

### Data Availability

Pretrained models, usage examples, and documentation of our downloadable python module can be found here (https://github.com/j-fife/clintegrate/). Precomputed feature vectors for all SNVs in the analyzed genes are also available in the repository.

## Data Availability

Pretrained models, usage examples, and documentation of our downloadable python module can be found online at https://github.com/j-fife/clintegrate/ .

## Acknowledgments

We are indebted to the UK Biobank and its participants who provided biological samples and data for this analysis. Work was performed under UK Biobank application #7089 and Mass General Brigham IRB protocol 2020P002093. We are also grateful for advice and assistance from Harvard Catalyst, Dr. Shamil Sunyaev, and Dr. Richard Sherwood.

---

**Supplementary Figure 1:**
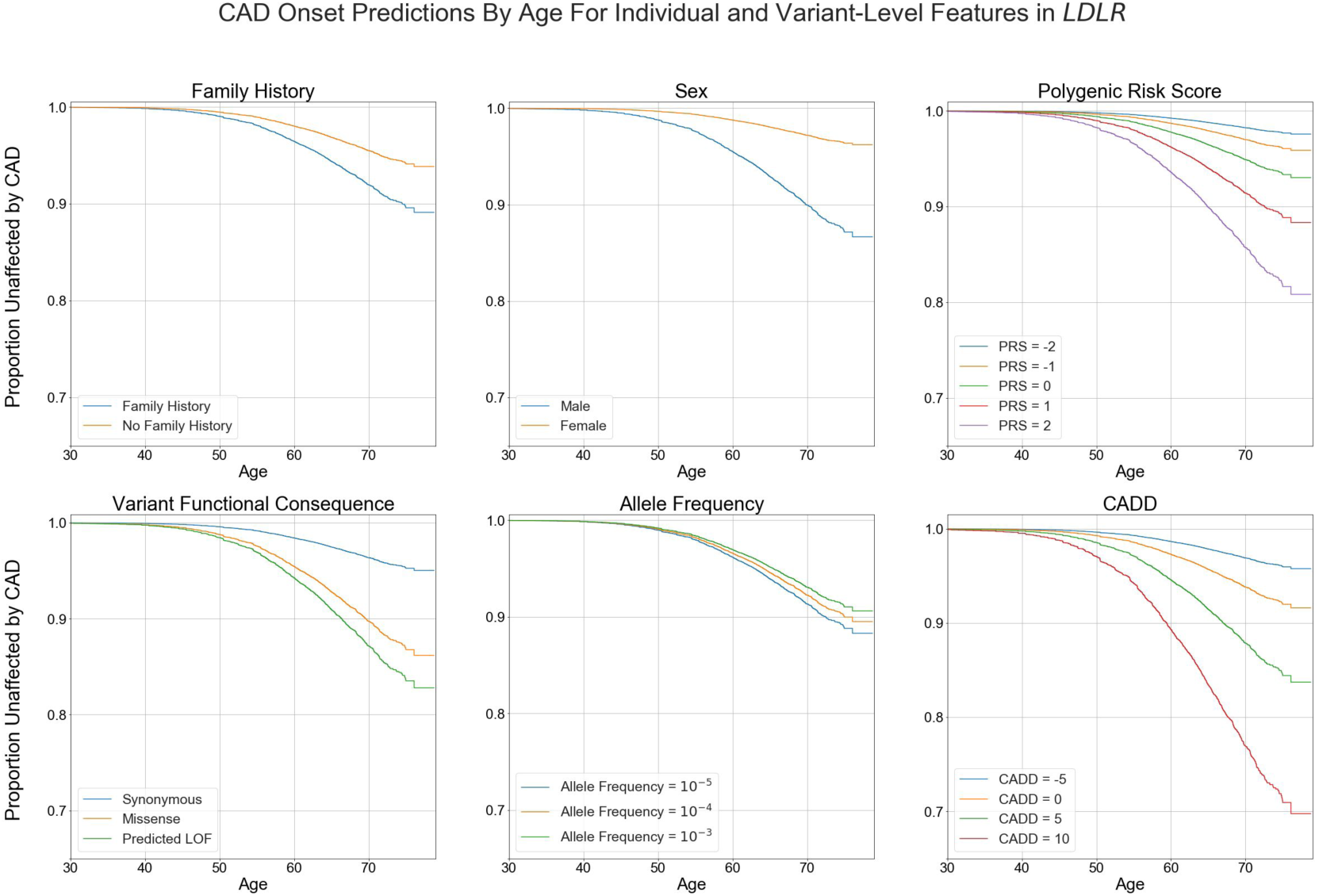
Univariate regressions of individual-level or variant-level predictive features for incidence of CAD, by age. Cox proportional hazards model fit using UKB patients to make predictions of CAD risk for patients with variants in *LDLR*. The model is fit using data for individual-level risk, including patient sex, family history of CAD, and polygenic risk score for CAD, as well as variant-level characteristics. All patient data are age-censored, using age at enrollment for prevalent CAD or time of incidence during the study period. Predictions can then be personalized using each individual’s variant-level and individual-level characteristics and age.

**Supplementary Figure 2:**
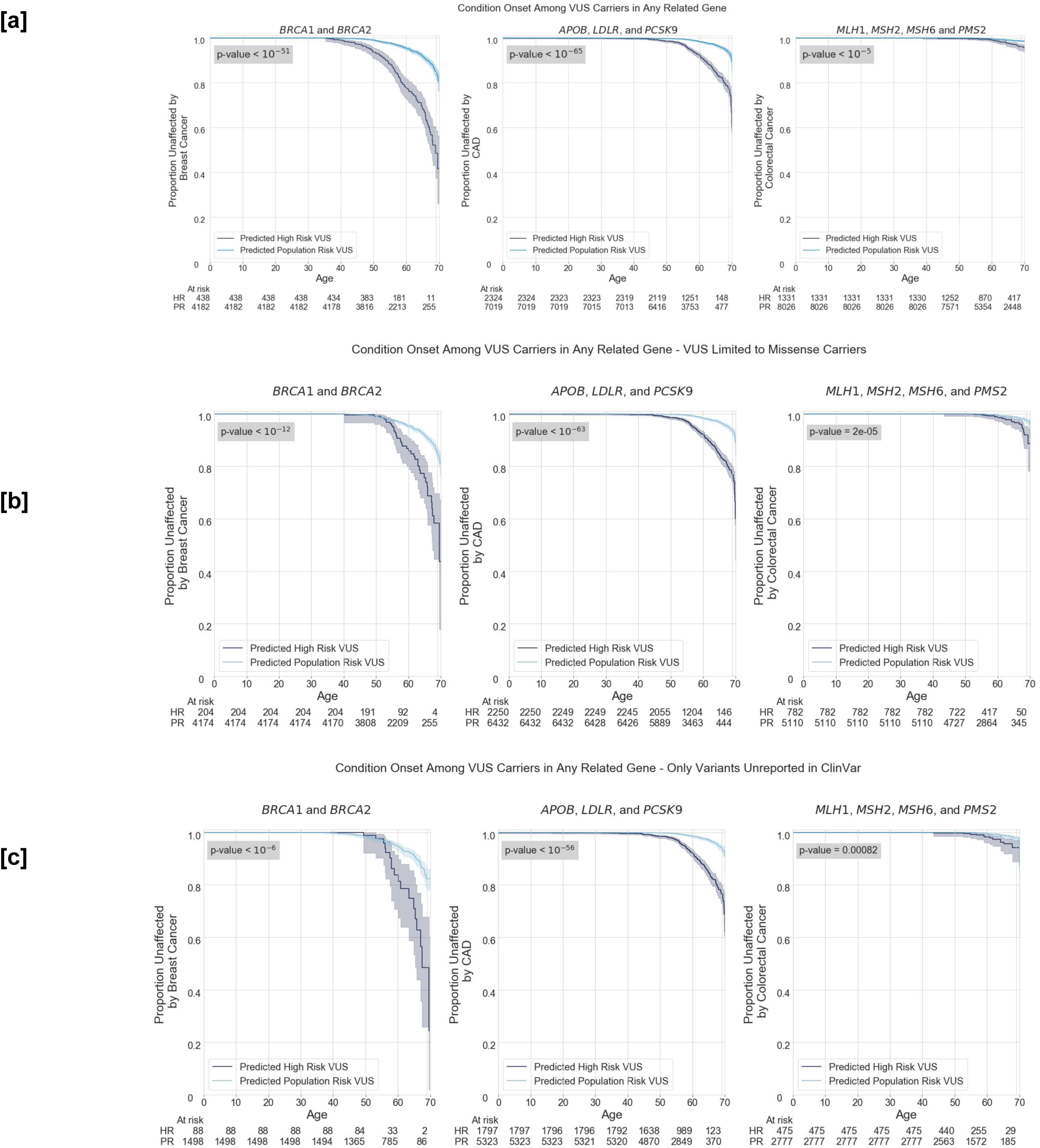
**[a]** We find significant differences in risk for breast cancer (BC), colorectal cancer (CRC), and coronary artery disease (CAD) among VUS carriers. Among those we classify as high risk (HR), and those we classify as low risk (LR) we find substantial risk ratios: RR=3.71 [3.53, 3.90] for BC, RR=4.71 [4.50, 4.92] for CAD, and RR=2.65 [2.15, 3.14] for CRC **[b]** Results are still significant when restricting to only individuals with missense VUSs: RR=2.90 [2.60, 3.20] for BC, RR = 2.58 [1.96, 3.21] for CRC, RR=4.52 [4.31, 4.73] for CAD, **[c]** Results are still significant when restricting to only those with variants unreported in ClinVar: RR=2.71 [2.26, 3.16] for breast cancer, RR=3.75 [4.26, 3.25] for CAD, RR = 3.32 [2.56, 4.07] for colorectal cancer.

**Supplementary Figure 3:**
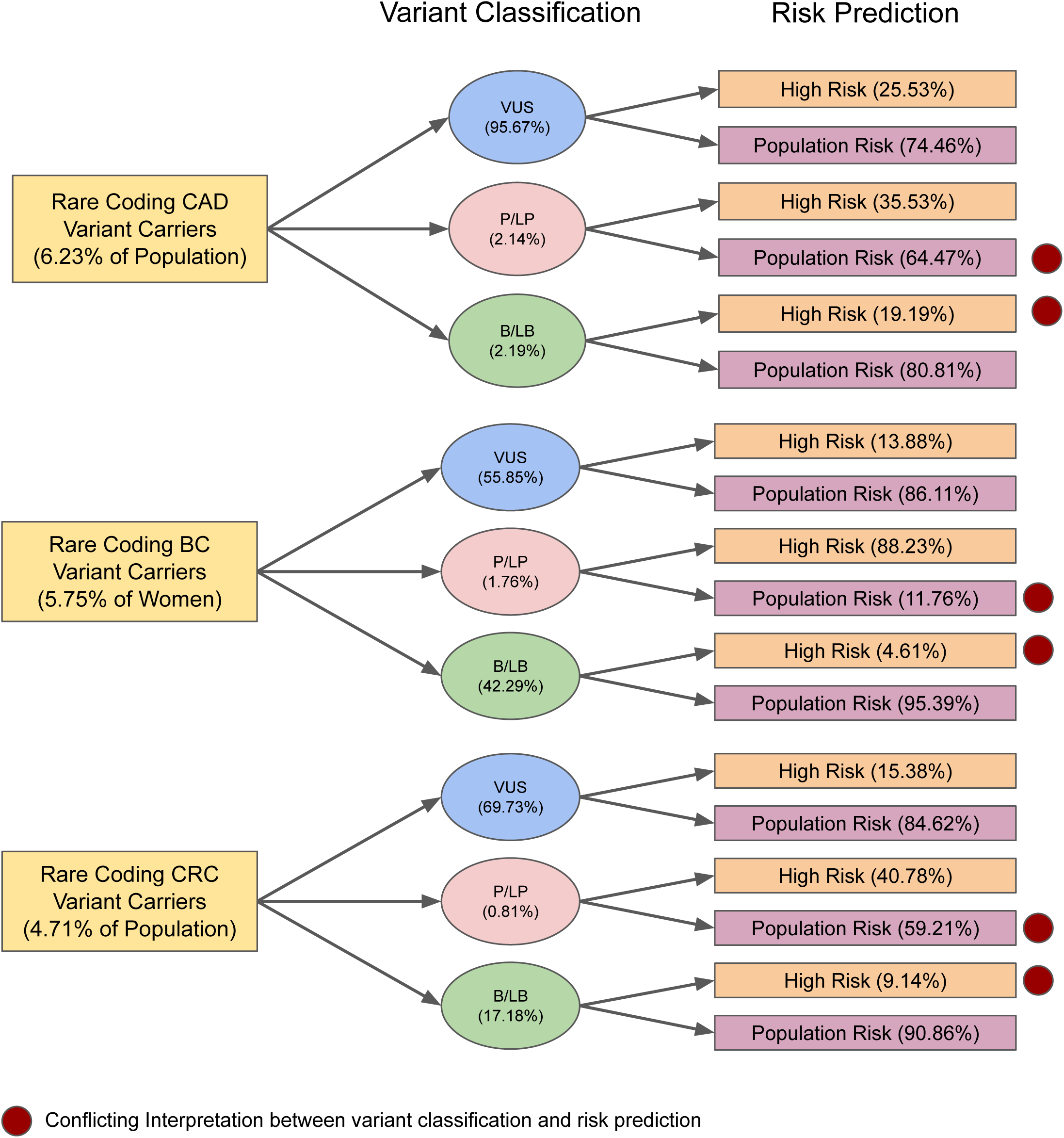
Rates of variant classifications and carrier risk predictions in the UK Biobank population. Pathogenic/Likely Pathogenic (P/LP) classifications were generated by the Mass General Brigham Lab for Molecular Medicine. Benign/Likely Benign (B/LB) classifications are drawn from ClinVar. The remaining rare non-synonymous variants are classified as Variants of Uncertain Significance (VUS). For each disorder, we find significant numbers of carriers who have discordant risk predictions from prior variant classifications.

**Supplementary Figure 4:**
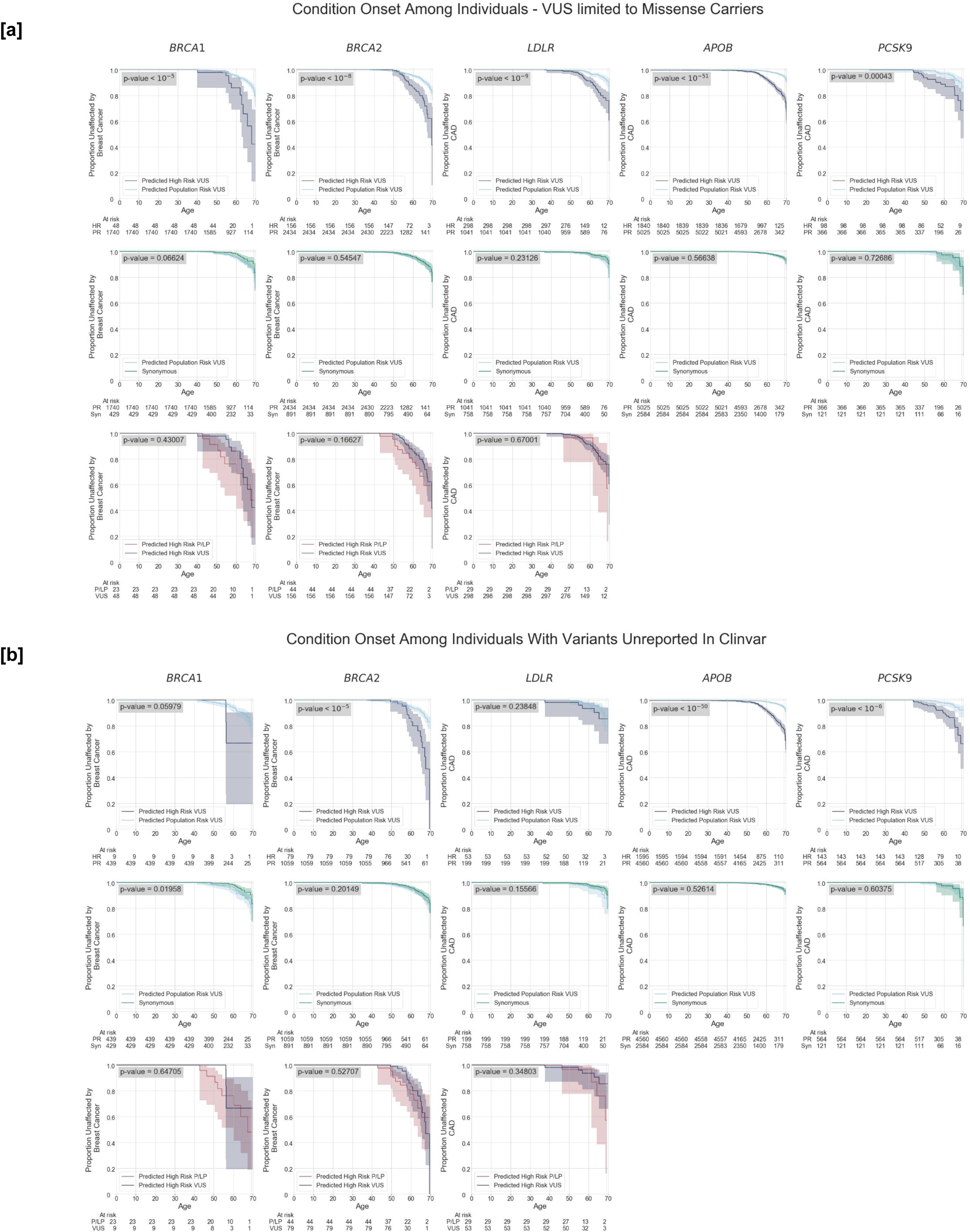
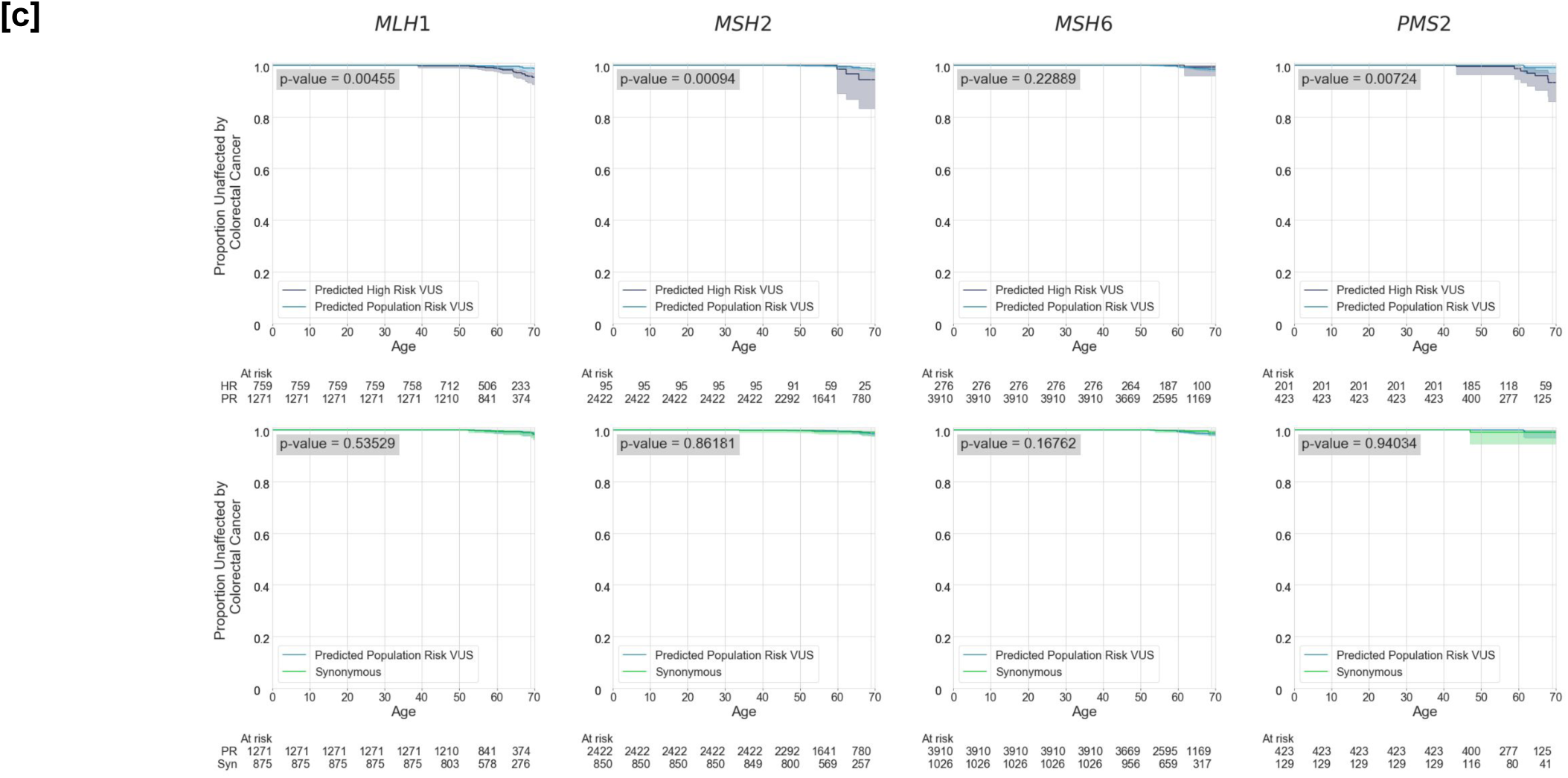
**[a]** Risk thresholds separate individuals with high and low clinical risk in all five genes examined (p < 10^-6^ in all cases) when restricting to missense VUSs. Additionally, we see no significant difference in risk when comparing individuals with VUS predicted to have population risk and individuals with synonymous variants. Further, those individuals with lab-classified P/LP variants predicted to have high risk did not develop disorders at higher rates than those with VUS in the same score range. **[b]** Many comparisons still reach significant thresholds when limiting to VUSs previously unreported in ClinVar despite much smaller sample sizes in some genes. **[c]** Comparisons from Figure 2A in colorectal cancer genes. Notably no genes show significance when comparing low risk VUS carriers to synonymous carriers.

**Supplementary Figure 5:**
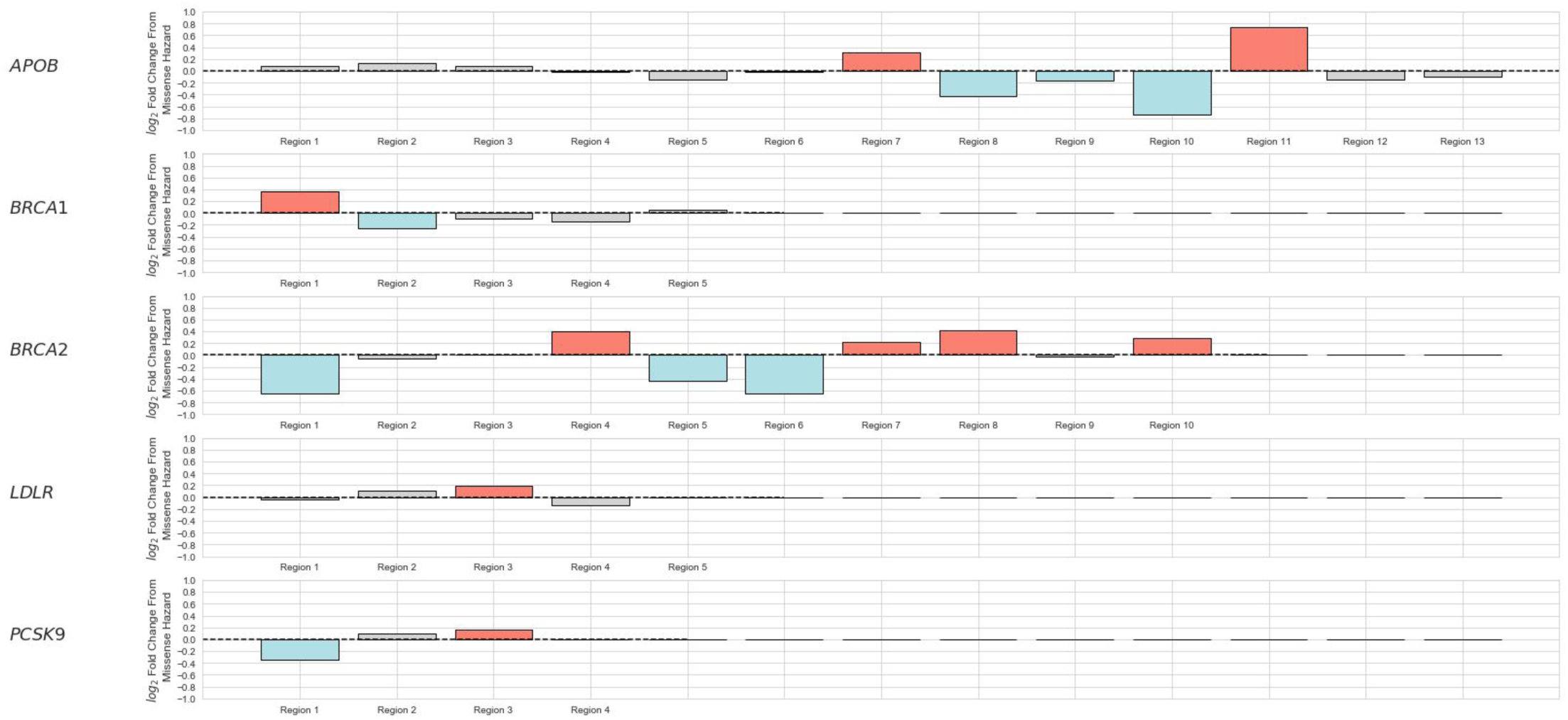
Each gene carries a differing level of risk from presence of a missense variant (**Supplementary Table 1**). We define regions of high and low risk as relative to each gene’s overall missense risk. Using univariate regressions on regions we define high risk as being above 1.1 log_2_ Fold Change from missense risk, and low risk as being below 0.90 log_2_ Fold Change from missense risk. Red bars indicate above 1.1 LFC, blue bars indicate below 0.9 LFC.

**Supplementary Figure 6:**
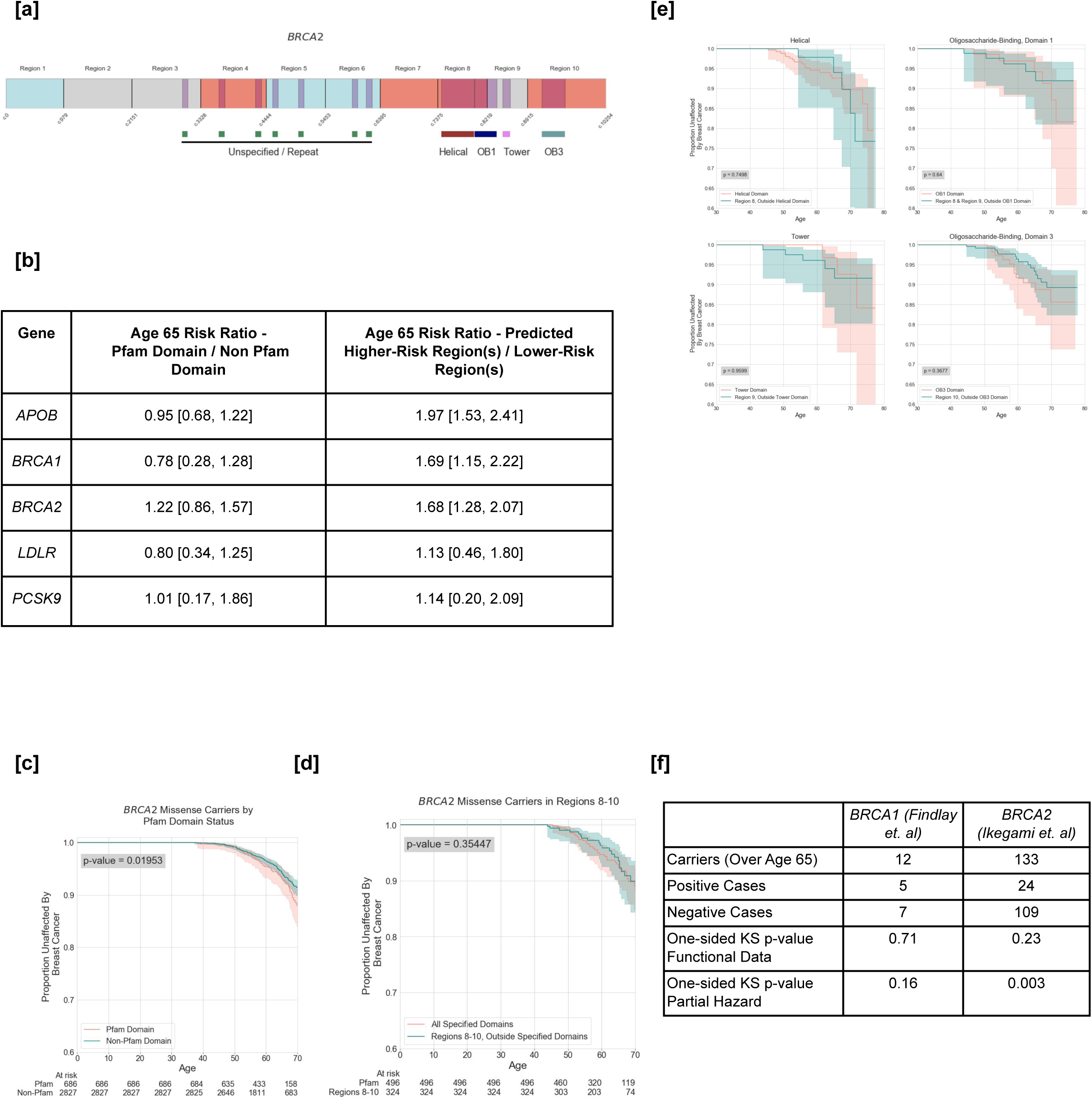
Comparison of regional boundaries to Pfam protein domains. **[a]** Schematic of the *BRCA2* transcript with Pfam protein domains (purple), predicted higher-risk regions (HRRs) (red), and lower-risk regions (LRR) (blue). **[b]** In the five genes analyzed, regional boundaries provide superior relative risks compared with Pfam protein domain boundaries. **[c]** Individuals with *BRCA2* variants within Pfam domains carry elevated risk for breast cancer (logrank p=0.0195). **[d]** The elevated-risk associated with Pfam domains is not limited to the domain itself. Specified Pfam domains partially overlap with predicted HRRs (Regions 8-10). We find no significant difference in breast cancer risk among individuals with variants within specified Pfam domains vs. individuals with variants outside those domains within Regions 8-10. **[e]** *BRCA2* Pfam domains overlap with our partitioned regions 8-10. In addition to few carriers in these domains, there are no significant differences in breast cancer onset among those with variants in the underlying regions but outside of Pfam domains. Individuals included in cyan KM curves have variants outside of all Pfam domains, not solely that domain specified in the comparison. **[f]** Functional data from *BRCA1* and *BRCA2* screens cover variants we observe in the UKB female population. These functional datasets, however, do not outperform the risk-predictions we assign these carriers.

**Supplementary Figure 7:**
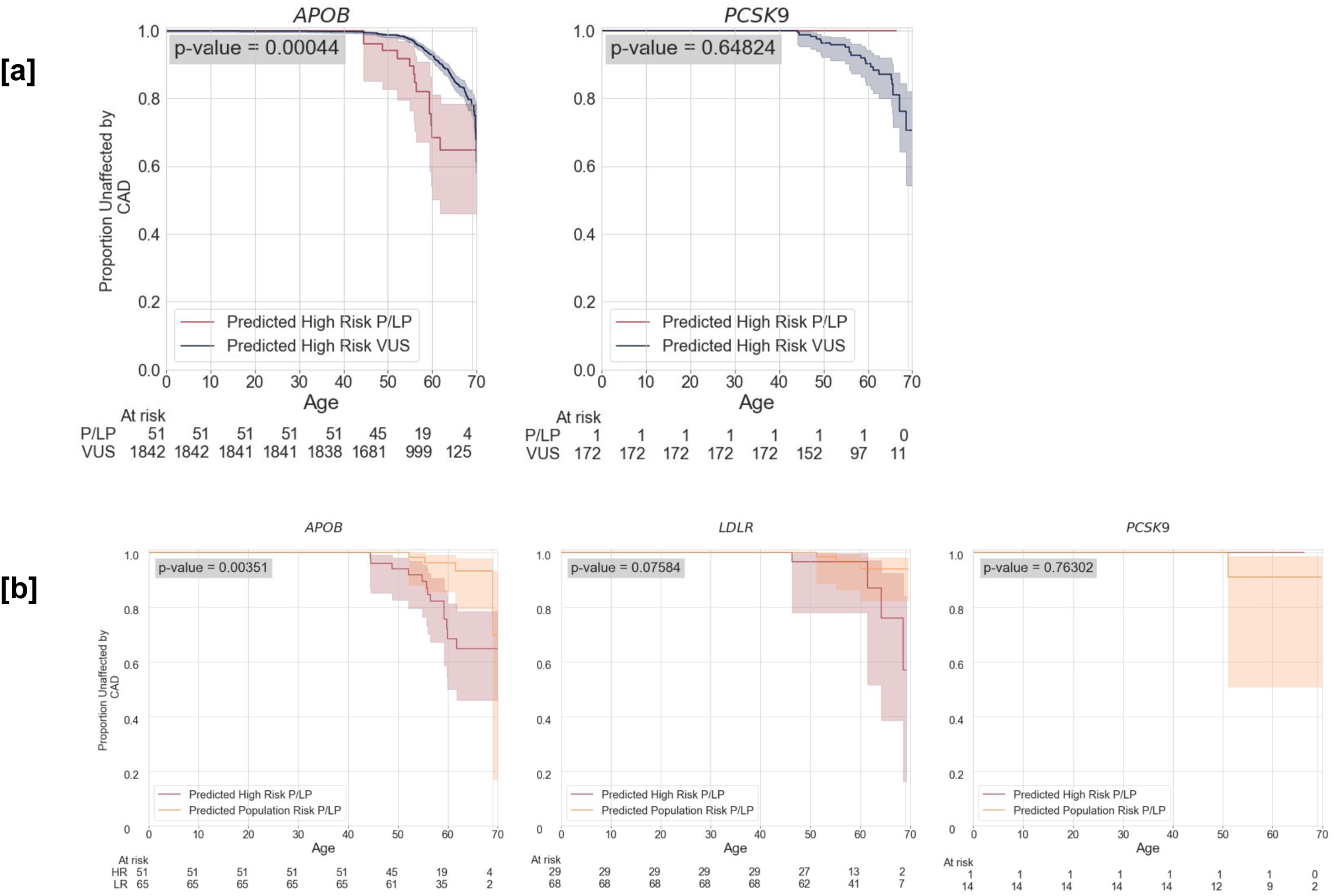
Classifying riak across variant annotations in genes with little statistical power or variant diversity. **[a]** In *APOB,* we analyze only one P/LP missense variant (2-21006288-C-T p.Arg3527Gln), which was highly penetrant for coronary artery disease (CAD) risk. Unsurprisingly, carriers of this variant develop CAD at higher rates than high-risk VUS carriers. Of 116 carriers, we predict 51 to be at high risk and 65 to be low risk. In *PCSK9,* among the 15 P/LP carriers only one individual developed CAD. 14/15 of these individuals were predicted to be population-risk, including the one individual who developed CAD. **[b]** Despite this single *APOB* variant being highly linked to CAD, we still find those we predict as high-risk develop CAD at a higher rate (RR=3.79 [2.68, 4.90] logrank p=0.00035) than those we predict as population-risk. In *LDLR,* we see similar outcomes with those predicted to be high-risk developing CAD at higher rates than those we predict as population-risk (RR=3.27 [1.95, 4.60] logrank p=0.076). In *PCSK9*, only 15 individuals carry known pathogenic variants, which may limit our ability to distinguish between HR and LR carriers of pathogenic variants.

**Supplementary Table 1:** We fit a Cox Proportional Hazards model on predictive features in each gene separately. We find varying levels of risk associated with variant-level features (e.g.,CADD, GERP) as well as varying levels of risk associated with carrying variants in each gene whether missense or predicted LOF (frameshift, splice-site, nonsense). Entries labeled N/A were caused by lack of model convergence when regressing on that feature due to lack of differential clinical outcomes among groups with and without that feature. In the case of *BRCA1* and *BRCA2,* sex was not used as a feature as predictive models include only women. Slight differences in individual-level features in genes related to the same condition (family history, sex, PRS) are due to exclusions of patients with coding variants in other associatd genes (see Methods) leading to a different set of patients for each gene model.

| Gene | Condition | CADD $\geq 5.0$<br>(95% CI) | GERP $\geq 4.0$<br>(95% CI) | phyloP $\geq 3.0$<br>(95% CI) | Allele Frequency<br>$\leq 10^{-4}$ (95% CI) |
| --- | --- | --- | --- | --- | --- |
| *BRCA1* | Breast Cancer | 7.71 [5.32, 11.17]*** | 1.3 [0.96, 1.76] | 1.26 [0.84, 1.88] | 1.22 [0.97, 1.53] |
| *BRCA2* | Breast Cancer | 5.48 [4.36, 6.88]*** | 1.06 [0.82, 1.38] | 1.17 [0.89, 1.54] | 1.44 [1.24, 1.66]*** |
| *APOB* | Coronary Artery Disease | 0.36 [0.14, 0.96]* | 1.17 [1.0, 1.37]* | 1.17 [0.98, 1.41] | 0.89 [0.77, 1.03] |
| *LDLR* | Coronary Artery Disease | 6.2 [2.78, 13.81]*** | 1.5 [1.12, 2.02]* | 1.53 [1.07, 2.17]* | 1.4 [1.08, 1.82]* |
| *PCSK9* | Coronary Artery Disease | 2.0 [0.5, 8.0] | 1.79 [1.06, 3.02]* | 1.36 [1.0, 1.86] | 1.18 [0.84, 1.65] |
| *MLH1* | Colorectal Cancer | N/A | 1.74 [1.26, 2.41]** | 1.73 [1.25, 2.4]** | 2.54 [1.74, 3.71]*** |
| *MSH2* | Colorectal Cancer | N/A | 1.07 [0.73, 1.58] | 1.03 [0.69, 1.52] | 0.8 [0.45, 1.41] |
| *MSH6* | Colorectal Cancer | 1.15 [0.62, 2.14] | 0.99 [0.63, 1.55] | 0.84 [0.59, 1.19] | 0.98 [0.69, 1.4] |
| *PMS2* | Colorectal Cancer | 3.77 [0.94, 15.07] | 0.74 [0.28, 1.97] | 0.86 [0.36, 2.08] | 1.16 [0.43, 3.08] |

**Supplementary Table 1:** We fit a Cox Proportional Hazards model on predictive features in each gene separately.
| Gene | Condition | Family History | PRS $\geq 2.5$<br>(95% CI) | Sex=Male | Missense | Predicted LoF |
| --- | --- | --- | --- | --- | --- | --- |
| *BRCA1* | Breast Cancer | 1.8 [1.68, 1.92]*** | 2.5 [2.19, 2.86]*** | N/A | 0.97 [0.82, 1.16] | 7.31 [5.07, 10.52]*** |
| *BRCA2* | Breast Cancer | 1.81 [1.7, 1.93]*** | 2.52 [2.21, 2.87]*** | N/A | 0.99 [0.86, 1.15] | 5.31 [4.24, 6.65]*** |
| *APOB* | Coronary Artery Disease | 1.77 [1.69, 1.85]*** | 3.29 [2.82, 3.83]*** | 4.86 [4.59, 5.14]*** | 1.07 [0.96, 1.19] | 0.34 [0.13, 0.91]* |
| *LDLR* | Coronary Artery Disease | 1.78 [1.69, 1.87]*** | 3.22 [2.74, 3.79]*** | 5.42 [5.09, 5.77]*** | 1.32 [1.07, 1.63]* | 6.61 [2.97, 14.72]*** |
| *PCSK9* | Coronary Artery Disease | 1.75 [1.67, 1.84]*** | 3.2 [2.73, 3.75]*** | 4.88 [4.6, 5.17]*** | 1.62 [1.17, 2.25]** | 2.26 [0.57, 9.04] |
| *MLH1* | Colorectal Cancer | 1.8 [1.6, 2.01]*** | 1.26 [0.75, 2.13] | 1.38 [1.25, 1.51]*** | 1.74 [1.22, 2.48]** | N/A |
| *MSH2* | Colorectal Cancer | 1.78 [1.59, 2.0]*** | 1.18 [0.68, 2.03] | 1.37 [1.25, 1.5]*** | 0.78 [0.49, 1.24] | 9.49 [4.26, 21.15]*** |
| *MSH6* | Colorectal Cancer | 1.77 [1.58, 1.99]*** | 1.17 [0.68, 2.01] | 1.37 [1.25, 1.5]*** | 1.16 [0.83, 1.62] | 1.15 [0.62, 2.14] |
| *PMS2* | Colorectal Cancer | 1.76 [1.57, 1.97]*** | 1.19 [0.69, 2.06] | 1.36 [1.24, 1.49]*** | 1.2 [0.54, 2.67] | 3.77 [0.94, 15.07] |

**Supplementary Table 2:** **[a]** We see individuals with Pathogenic or Likely Pathogenic variants receiving higher predicted hazard scores than individuals with Variants of Uncertain Significance (VUS). While by and large these annotations are recapitulated through our predictions, these classifications alone are not sufficient at separating those with increased risk of the associated condition. **[b]** Along with higher distributions of scores overall, Individuals with VUS received a lower predicted hazard score on average.

| Gene | VUS Predictions vs. P/LP Predictions (Mann Whitney U p-value) |
| --- | --- |
| BRCA1 | 5.11x10<sup>-17</sup> |
| BRCA2 | 9.10x10<sup>-28</sup> |
| LDLR | 1.57x10<sup>-4</sup> |

**Supplementary Table 2:** [a] We see individuals with Pathogenic or Likely Pathogenic variants receiving higher predicted hazard scores than individuals with Variants of Uncertain Significance (VUS).
| Gene | Variant of Uncertain Significance (VUS) | Pathogenic / Likely Pathogenic (P/LP) |
| --- | --- | --- |
| BRCA1 | 1.39 [1.3, 1.47] | 7.06 [5.1, 9.01] |
| BRCA2 | 1.51 [1.45, 1.57] | 6.51 [5.19, 7.83] |
| LDLR | 2.18 [2.04, 2.33] | 3.43 [2.3, 4.57] |

**Supplementary Table 3:**
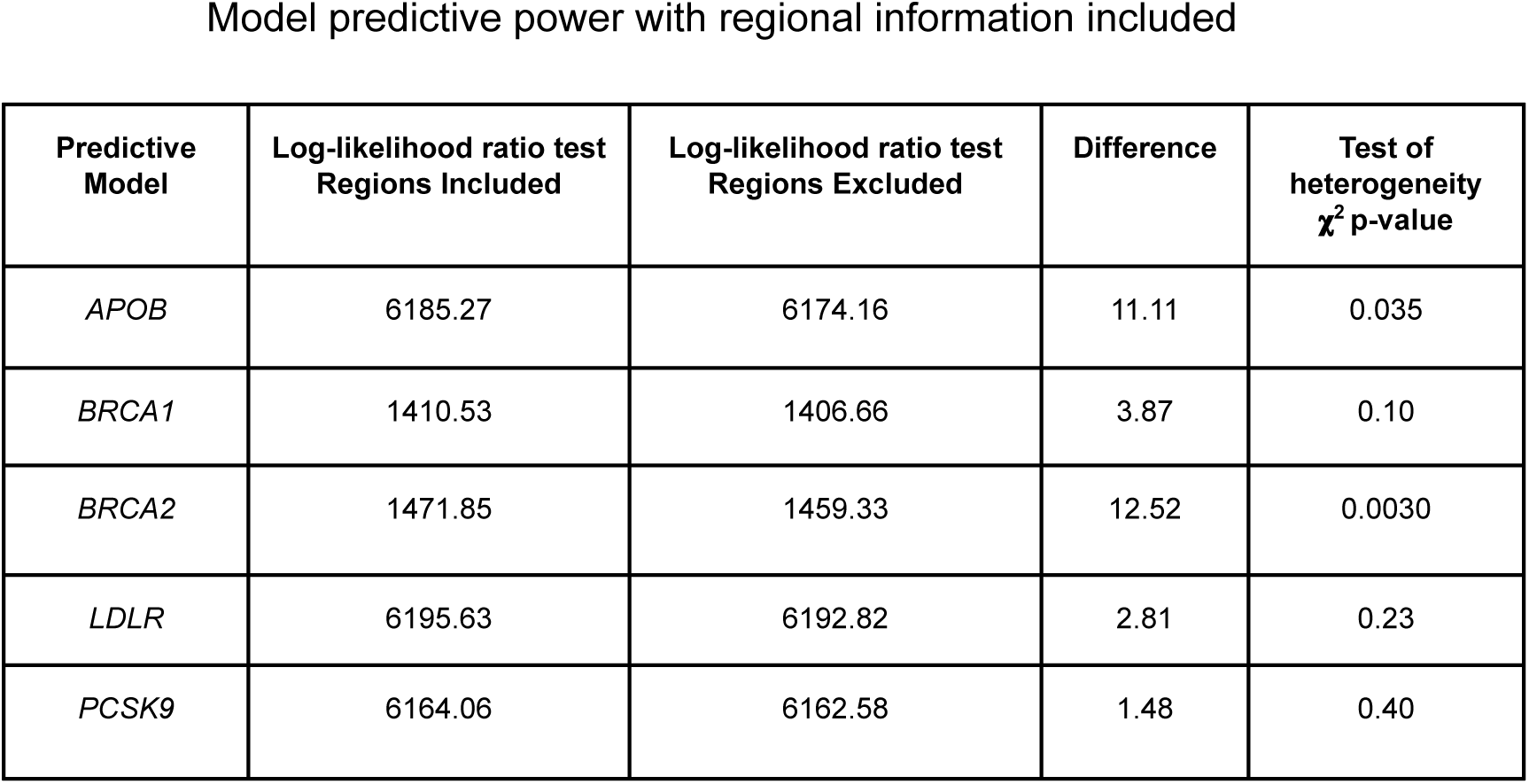
We fit predictive models with and without regional definitions as parameters. In genes where significant logrank p-value was not reached through a high and low risk regional comparison (*BRCA1, LDLR*), test of heterogeneity *χ*^2^ p-value also significant. Log-Likelihood ratio test also indicated better fit of the predictive models in all five genes examined.

**Supplementary Table 4:**
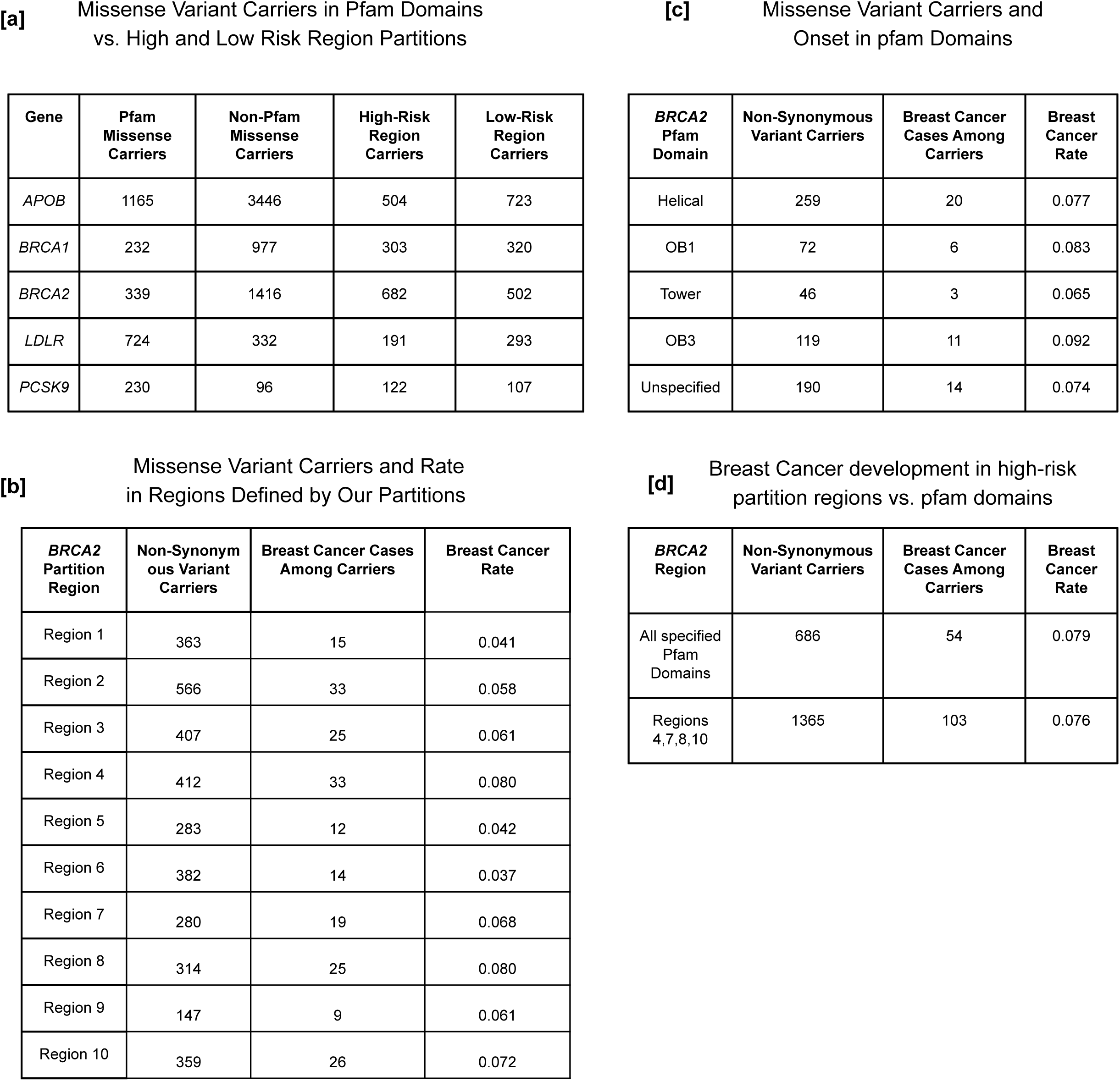
In the five genes analyzed, Pfam domains in *BRCA2* showed the most signal in terms of individuals with variants in these domains, and signal measured by the Cox Model. Even then, however, cases remained sparse, and onset in these individuals remained unreliable to the extent that statistical significance was not reached from examining Pfam domains on their own.

| Gene | Pfam Missense Carriers | Non-Pfam Missense Carriers | High-Risk Region Carriers | Low-Risk Region Carriers |
| --- | --- | --- | --- | --- |
| APOB | 1165 | 3446 | 504 | 723 |
| BRCA1 | 232 | 977 | 303 | 320 |
| BRCA2 | 339 | 1416 | 682 | 502 |
| LDLR | 724 | 332 | 191 | 293 |
| PCSK9 | 230 | 96 | 122 | 107 |

[c] Missense Variant Carriers and Onset in pfam Domains
| BRCA2 Pfam Domain | Non-Synonymous Variant Carriers | Breast Cancer Cases Among Carriers | Breast Cancer Rate |
| --- | --- | --- | --- |
| Helical | 259 | 20 | 0.077 |
| OB1 | 72 | 6 | 0.083 |
| Tower | 46 | 3 | 0.065 |
| OB3 | 119 | 11 | 0.092 |
| Unspecified | 190 | 14 | 0.074 |

[b] Missense Variant Carriers and Rate in Regions Defined by Our Partitions
| BRCA2 Partition Region | Non-Synonymous Variant Carriers | Breast Cancer Cases Among Carriers | Breast Cancer Rate |
| --- | --- | --- | --- |
| Region 1 | 363 | 15 | 0.041 |
| Region 2 | 566 | 33 | 0.058 |
| Region 3 | 407 | 25 | 0.061 |
| Region 4 | 412 | 33 | 0.080 |
| Region 5 | 283 | 12 | 0.042 |
| Region 6 | 382 | 14 | 0.037 |
| Region 7 | 280 | 19 | 0.068 |
| Region 8 | 314 | 25 | 0.080 |
| Region 9 | 147 | 9 | 0.061 |
| Region 10 | 359 | 26 | 0.072 |

**Supplementary Table 4:** In the five genes analyzed, Pfam domains in *BRCA2* showed the most signal in terms of individuals with variants in these domains, and signal measured by the Cox Model.
| BRCA2 Region | Non-Synonymous Variant Carriers | Breast Cancer Cases Among Carriers | Breast Cancer Rate |
| --- | --- | --- | --- |
| All specified Pfam Domains | 686 | 54 | 0.079 |
| Regions 4,7,8,10 | 1365 | 103 | 0.076 |

**Supplementary Table 5:** Testing model consistency when making predictions on a set of participants outside of the UKB. Here we restrict to individuals with non-synonymous variants not previously observed in the UKB. **[a]** Despite considerably lower statistical power than previous comparisons with no filter on observed variants, average predicted scores are significantly different by disease status (Mann-Whitney U p-value < 0.05) in some models. **[b]** Validation of classification thresholds in partitioning high and low-risk individuals in five genes with their related phenotypes when restricting to variants not previously observed in UKB participants.

| Gene | Phenotype | Individuals with Phenotype | Individuals without Phenotype | Average Score among Positive Phenotypes (95% CI) | Average Score among Negative Phenotypes (95% CI) | Mann-Whitney U p-value |
| --- | --- | --- | --- | --- | --- | --- |
| *APOB* | CAD | 25 | 137 | 2.26 [1.17, 3.35] | 1.43 [1.17, 1.7] | 0.11 |
| *LDLR* | CAD | 22 | 28 | 5.53 [3.27, 7.78] | 2.08 [1.04, 3.13] | 0.0010 |
| *PCSK9* | CAD | 2 | 16 | 3.76 [-33.6, 41.13] | 2.17 [1.03, 3.3] | 0.22 |
| *BRCA1* | BC | 1 | 14 | 7.15 [N/A, N/A] | 0.81 [0.61, 1.02] | 0.066 |
| *BRCA2* | BC | 4 | 52 | 1.53 [-0.44, 3.51] | 1.24 [1.01, 1.47] | 0.42 |

**Supplementary Table 5:** Testing model consistency when making predictions on a set of participants
| Gene | Phenotype | Partial Hazard Cutoff | Development Among Individuals Above Cutoff | Development Among Individuals Below Cutoff | Age 65 Risk Ratio (95% CI) | Phenotype Development $\chi^2$ p-value |
| --- | --- | --- | --- | --- | --- | --- |
| *APOB* | CAD | 2.1 | 9 / 38 | 16 / 124 | 1.84 [1.1, 2.57] | 0.18 |
| *LDLR* | CAD | 2.8 | 12 / 17 | 10 / 33 | 2.33 [1.73, 2.93] | 0.016 |
| *PCSK9* | CAD | 3.0 | 1 / 5 | 1 / 13 | 2.6 [0.03, 5.17] | 0.93 |
| *BRCA1* | BC | 2.5 | 1 / 1 | 0 / 14 | N/A | 0.072 |
| *BRCA2* | BC | 2.3 | 1 / 4 | 3 / 52 | 4.33 [2.31, 6.36] | 0.67 |

**Supplementary Table 6:** Overall incidence of the validated conditions in the Geisinger MyCode biobank and in the UKB. Breast cancer development rates are limited to only female participants. Development rates are relative to the size of the overall biobank populations respectively.

| Phenotype | Development By Age 60 | Development Within Lifetime |
| --- | --- | --- |
| CAD | 6.81% | 17.24% |
| Breast Cancer | 1.27% | 2.50% |

| Phenotype | Development By Age 60 | Development Within Lifetime |
| --- | --- | --- |
| CAD | 3.81% | 4.76% |
| Breast Cancer | 4.74% | 5.89% |

**Regeneron Genetics Center and Geisinger DiscovEHR Collaboration Banner Author List and Contribution Statements**

All authors/contributors are listed in alphabetical order.

**Geisinger personnel**

Lance J. Adams, Jackie Blank, Dale Bodian, Derek Boris, Adam Buchanan, David J. Carey, Ryan D. Colonie, F. Daniel Davis, Dustin N. Hartzel, Melissa Kelly, H. Lester Kirchner, Joseph B. Leader, J. Neil Manus, Christa L. Martin, Michelle Meyer, Tooraj Mirshahi, Matthew Oetjens, Christopher Still, Natasha Strande, Amy Sturm, Marc Williams

**RGC Management and Leadership Team**

Goncalo Abecasis, Aris Baras, Michael Cantor, Giovanni Coppola, Andrew Deubler, Aris Economides, Luca A. Lotta, John D. Overton, Jeffrey G. Reid, Alan Shuldiner, Katia Karalis and Katherine Siminovitch

Contribution: All authors contributed to securing funding, study design and oversight. All authors reviewed the final version of the manuscript.

**Sequencing and Lab Operations**

Christina Beechert, Caitlin Forsythe, Erin D. Fuller, Zhenhua Gu, Michael Lattari, Alexander Lopez, John D. Overton, Thomas D. Schleicher, Maria Sotiropoulos Padilla, Louis Widom, Sarah E. Wolf, Manasi Pradhan, Kia Manoochehri, Ricardo H. Ulloa.

Contribution: C.B., C.F., A.L., and J.D.O. performed and are responsible for sample genotyping. C.B, C.F., E.D.F., M.L., M.S.P., L.W., S.E.W., A.L., and J.D.O. performed and are responsible for exome sequencing. T.D.S., Z.G., A.L., and J.D.O. conceived and are responsible for laboratory automation. M.S.P., K.M., R.U., and J.D.O are responsible for sample tracking and the library information management system.

**Genome Informatics**

Xiaodong Bai, Suganthi Balasubramanian, Boris Boutkov, Gisu Eom, Lukas Habegger, Alicia Hawes, Shareef Khalid, Olga Krasheninina, Rouel Lanche, Adam J. Mansfield, Evan K. Maxwell, Mona Nafde, Sean O’Keeffe, Max Orelus, Razvan Panea, Tommy Polanco, Ayesha Rasool, Jeffrey G. Reid, William Salerno, Jeffrey C. Staples,

Contribution: X.B., A.H., O.K., A.M., S.O., R.P., T.P., A.R., W.S. and J.G.R. performed and are responsible for the compute logistics, analysis and infrastructure needed to produce exome and genotype data. G.E., M.O., M.N. and J.G.R. provided compute infrastructure development and operational support. S.B., S.K., and J.G.R. provide variant and gene annotations and their functional interpretation of variants. E.M., J.S., R.L., B.B., A.B., L.H., J.G.R. conceived and are responsible for creating, developing, and deploying analysis platforms and computational methods for analyzing genomic data.

**Pharmacogenomics Genetics:**

Charles Paulding

Contribution: Contributed to the review process for the final version of the manuscript

**Research Program Management**

Marcus B. Jones, Jason Mighty, and Lyndon J. Mitnaul

Contribution: All authors contributed to the management and coordination of all research activities, planning and execution. All authors contributed to the review process for the final version of the manuscript.

